# Whole genome sequence-based association analysis of African American individuals with bipolar disorder and schizophrenia

**DOI:** 10.1101/2024.12.27.24319111

**Authors:** Runjia Li, Sarah A. Gagliano Taliun, Kevin Liao, Matthew Flickinger, Janet L. Sobell, Giulio Genovese, Adam E. Locke, Rebeca Rothwell Chiu, Jonathon LeFaive, Jiongming Wang, Taylor Martins, Sinéad Chapman, Anna Neumann, Robert E. Handsaker, Donna K. Arnett, Kathleen C. Barnes, Eric Boerwinkle, David Braff, Brian E. Cade, Myriam Fornage, Richard A. Gibbs, Karin F. Hoth, Lifang Hou, Charles Kooperberg, Ruth J.F. Loos, Ginger A. Metcalf, Courtney G. Montgomery, Alanna C. Morrison, Zhaohui S. Qin, Susan Redline, Alexander P. Reiner, Stephen S. Rich, Jerome I. Rotter, Kent D. Taylor, Karine A. Viaud-Martinez, NHLBI Trans-Omics for Precision Medicine (TOPMed) Consortium, Genomic Psychiatry Cohort investigators, Tim B. Bigdeli, Stacey Gabriel, Sebastian Zollner, Albert V. Smith, Goncalo Abecasis, Steve McCarroll, Michele T. Pato, Carlos N. Pato, Michael Boehnke, James Knowles, Hyun Min Kang, Roel A. Ophoff, Jason Ernst, Laura J. Scott

## Abstract

In studies of individuals of primarily European genetic ancestry, common and low-frequency variants and rare coding variants have been found to be associated with the risk of bipolar disorder (BD) and schizophrenia (SZ). However, less is known for individuals of other genetic ancestries or the role of rare non-coding variants in BD and SZ risk. We performed whole genome sequencing of African American individuals: 1,598 with BD, 3,295 with SZ, and 2,651 unaffected controls (InPSYght study). We increased power by incorporating 14,812 jointly called psychiatrically unscreened ancestry-matched controls from the Trans-Omics for Precision Medicine (TOPMed) Program for a total of 17,463 controls. To identify variants and sets of variants associated with BD and/or SZ, we performed single-variant tests, gene-based tests for singleton protein truncating variants, and rare and low-frequency variant annotation-based tests with conservation and universal chromatin states and sliding windows. We found suggestive evidence of BD association with single-variants on chromosome 18 and of lower BD risk associated with rare and low-frequency variants on chromosome 11 in a region with multiple BD GWAS loci, using a sliding window approach. We also found that chromatin and conservation state tests can be used to detect differential calling of variants in controls sequenced at different centers and to assess the effectiveness of sequencing metric covariate adjustments. Our findings reinforce the need for continued whole genome sequencing in additional samples of African American individuals and more comprehensive functional annotation of non-coding variants.

## Introduction

Severe mental illnesses, including bipolar disorder (BD) and schizophrenia (SZ), are debilitating disorders affecting millions of people worldwide. BD and SZ encompass a wide range of shared symptoms including recurrent episodes of psychosis, large mood swings, depression, and cognitive impairment. Both disorders are significantly associated with risk for suicide and increased all-cause mortality rate^1^. Heritability estimates from family studies range from 60%-85% for BD^2^ and 60%-80% for SZ^3^. Notably, there is considerable overlap in the underlying genetics of BD and SZ^4^, and genetic correlations are estimated to be as high as 0.68 at the common variant level^5^. Uncovering the genetic factors contributing to these disorders could lead to a deeper understanding of disease etiology and improved treatment options.

Hundreds of independent susceptibility loci for BD and SZ have been identified through large-scale genome-wide association studies (GWAS) by focusing on common and low-frequency alleles ^6–14^. SZ case-control GWAS of European and East Asian ancestry individuals (Psychiatric Genomics Consortium, 76,755 individuals with SZ and 243,649 controls) identified 287 distinct loci, implicating genes associated with neurodevelopmental disorders and with brain-specific expression^13^. A BD GWAS of European ancestry (41,917 BD cases and 371,549 controls) identified 64 distinct loci, and significant enrichment of association signals within genes belonging to neuronal and synaptic pathways and targets for existing BD medications^4^. Of the BD-associated variants, 17 were also associated with SZ. For both the BD and SZ GWAS, measured common variants are estimated to account for a modest portion of disease heritability (18.6% for BD^4^ and 24% for SZ^15^).

Whole-exome sequencing (WES) detects coding variants across the allele frequency spectrum. The Schizophrenia Exome Sequencing Meta-Analysis (SCHEMA) consortium, produced a SZ case-control WES study (SCHEMA) of multi-ancestry individuals (24,248 SZ cases and 97,322 controls), which identified enrichment of ultra-rare coding variants in ten genes in individuals with SZ^16^; two of the genes were also implicated by common-variant GWAS^13^. A WES BD case-control study (BipEx) of European ancestry individuals (13,933 BD cases and 14,422 controls) found that, compared to controls, individuals with BD were enriched for ultra-rare protein truncating variants in constrained genes (probability of being loss-of-function intolerant (*p*LI) *≥* 0.9). When the BipEx and SCHEMA results were combined, *AKAP11* was identified as a risk gene^17^. These results identified ultra rare coding variants as contributing to BD and SZ risk and suggest overlap between BD and SZ risk at both the rare and common variant level.

Compared to common and exonic variants, less is known about the role of rare non-coding variants in SZ and BD. WGS allows detection of coding and non-coding variants across the allele frequency spectrum. Studies using WGS to investigate SZ or BD have generally been limited for non-coding variant analysis by sample sizes of no more than a few hundred individuals, often in family-based designs^18–24^. One SZ WGS case-control study of Swedish samples (1162 SZ cases and 936 controls) found association between SZ and structural variants at topologically associated domain boundaries but did not find significant differential burden of non-coding single nucleotide variants (SNVs) and insertion/deletion polymorphisms (indels) between SZ cases and controls across a variety of biological groupings^25^.

To date, genomic studies of psychiatric disorders, and many complex human diseases and traits, have overwhelmingly been composed of individuals of European genetic ancestry^26,27^. Although there has been progress in increasing the representation of ancestral backgrounds of individuals included in GWAS studies, notably in East Asians, the available data do not comprehensively represent individuals in the US or the world^28^. This impedes discovery of genes and mechanisms that might be uncovered from the broader spectrum variation across different ancestries. The use of European ancestry BD polygenic risk score (PRS) to predict disease risk across ancestries also has the potential to create health inequities. For example, PRS from European ancestry GWAS predict a much smaller proportion of disease in East Asian and African American ancestry samples than in European ancestry samples^4^. Increasingly, efforts are underway to assess the influence of genetic variation on complex traits in individuals of non-European ancestry ^29–31^. Genetic studies of mental health disorders that include WGS for individuals of diverse genetic ancestries will allow us to better address the disparities in diagnosis and treatment.

We examined the role of SNVs and short indels in BD and SZ susceptibility in African American individuals in a sample of 7544 individuals (with 1598 BD cases, 3295 SZ cases, and 2651 controls without SZ or BD) and an additional 14,812 phenotypically-unscreened ancestry-matched individuals from the NHLBI TOPMed program as external controls^31^. Overall, we found suggestive evidence of single-variant BD association on chromosome 18. We observed that chromatin and conservation state burden tests were a sensitive way to assess the comparability of sequencing/calling between two WGS sample sets. We also found suggestive evidence of association of a chromosome 11 region sliding window located among multiple previously reported BD GWAS loci^4^.

## Subjects and Methods

Figure 1 contains an overview of the study design, samples and analytical approaches.

**Figure 1.**
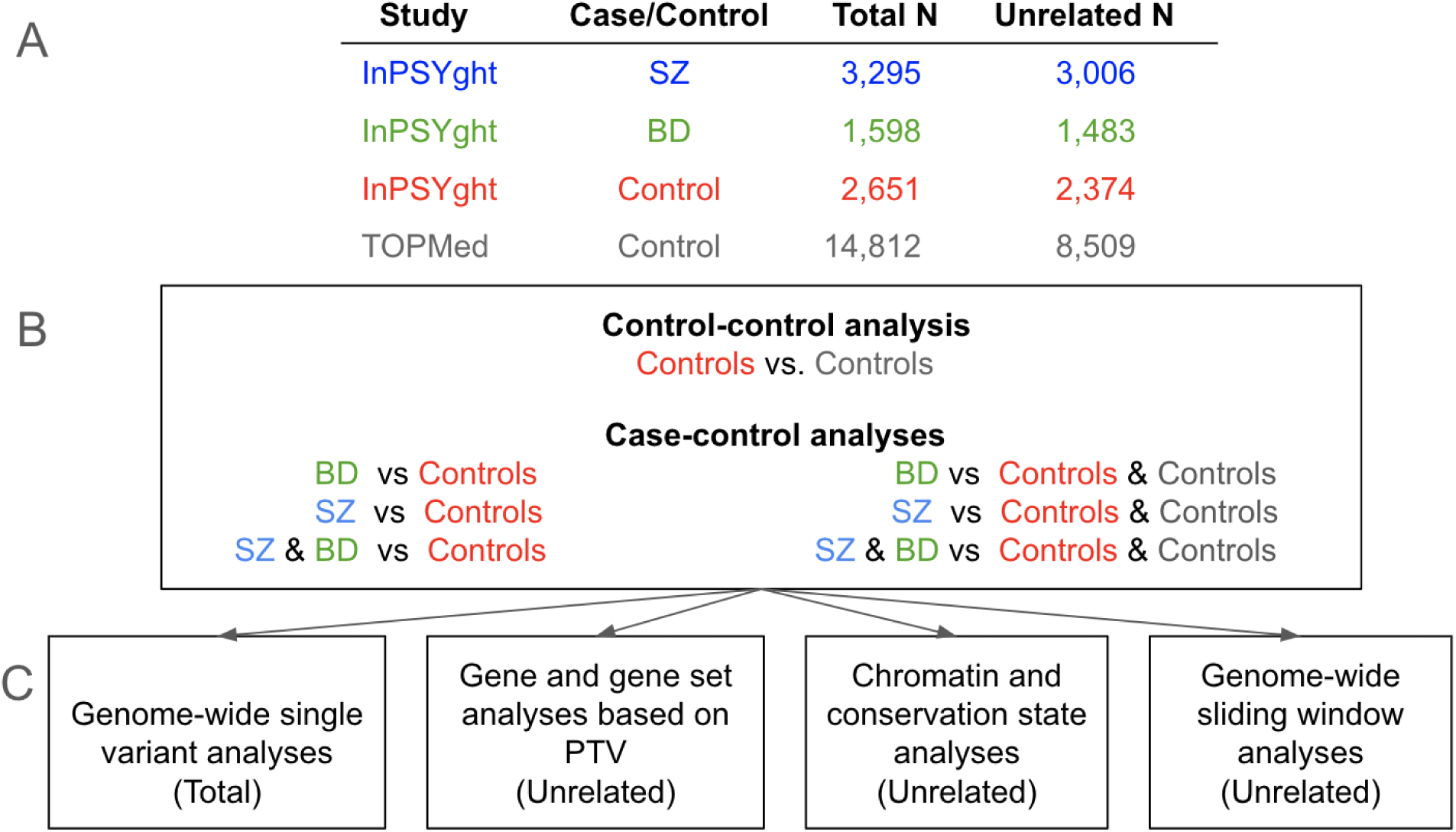
Study overview. (A) Number of total and of unrelated study participants for each case or control group; (B) Seven analysis groups: control/control and case/control; (C) Four analysis types (Total or Unrelated samples used in analysis); PTV: protein truncating variant

### InPSYght study sample

We performed deep whole-genome sequencing in a United States-based case-control study (the InPSYght study) of African American individuals as part of the Whole Genome Sequence for Psychiatric Disorders (WGSPD) consortium^32^. We use the term African American to denote individuals who self identified as African American on study forms; this term (which may have been one of a limited number of choices to describe African Ancestry) can include individuals who, among others, are descendants of enslaved individuals or are more recent immigrants to the US from African and other countries. The InPSYght study is composed of participants from the Genomic Psychiatry Cohort (GPC)^33,34^, Consortium on the Genetics of SZ (COGS)^35^, the Bipolar Genome Study (BIGS)^36^, Lithium treatment moderate dose use study (LiTMUS)^37^ and Systematic Treatment Enhancement Program for Bipolar Disorder (STEP-BD) studies^38^. All DNA samples were obtained from the NIMH Repository and Genomics Resource (**Table S1**). From these studies, we selected individuals with self-reported African/African American ancestry. We used ADMIXTURE^39^ to estimate the percent African genetic ancestry in each individual using study sample genotype array data and the 1000 Genomes^40^ ancestry super-populations: (Admixed Americans (AMR), African (AFR), European (EUR), South Asian (SAS), and East Asian (EAS)) as reference populations. We retained individuals with estimated percent global African genetic ancestry >25%. All individuals designated as cases fulfilled the DSM-IV criteria for SZ or BD. InPSYght controls (all from the GPC study) were included if they had no personal or family history of SZ or BD and if they did not have unipolar depression (from screening questions). Details of the recruitment strategies and instruments used for diagnosis are provided in the referenced publications for each cohort. All participants provided informed consent.

### TOPMed external controls

To increase statistical power to detect variants associated with BD and SZ, we selected as external controls previously whole genome sequenced self-identified African American individuals from the NHLBI Trans-Omics for Precision Medicine (TOPMed) project (hereafter referred to as TOPMed controls). TOPMed studies (case-control or family-based studies) are focused on heart, lung, blood and sleep disorders and inclusion was not predicated on information about mental health ^31^. We considered for inclusion individuals from 11 studies which agreed to the use of their samples as controls (**Table S2**). We considered for inclusion individuals with general research use (GRU) consent or consent for health/medical/biomedical purposes (HMB). Further inclusion/exclusion criteria are described below.

### Whole-genome sequencing of InPSYght and TOPMed individuals

#### InPSYght

Individuals were whole-genome sequenced (WGS; mean +/− SD depth 26.8 +/−5.5) in seven batches at the Broad Institute on an Illumina HiSeq X10 instrument. The first WGS batch (n=231 samples) was performed with PCR-based library preparation; the remaining batches were completed PCR free. All sequencing was paired end with 151 nucleotide read length. Individuals with SZ, with BD, and control individuals were included in each sequencing batch, except the PCR-based batch which did not include individuals with BD (**Table S3**). Case and internal control samples were intermixed within each batch to help avoid sub-batch effects.

#### TOPMed

Individuals were whole-genome sequenced at five sequencing centers as previously described^31^ (**Table S2**), using paired-end sequencing and read length of 150 base pairs. The 14,812 samples used as controls had a mean sequencing depth of 36.8 +/−4.7.

### Joint InPSYght and TOPMed variant discovery, genotype calling, and quality control

We used jointly called genotypes on human genome build version GRCh38 for bi-allelic SNVs and short indels from TOPMed Freeze 9^31^ using the GotCloud/vt pipeline^39^. We called genotypes for each individual on the autosomes and chromosome X on the across-individuals union of all variant sites. Genotypes for variants on the non-psuedo autosomal (nonPAR) region of the X chromosome were coded as 0 or 2 alleles for males, whereas those in the two pseudo autosomal regions (PAR1 and PAR2, respectively) were coded 0, 1, or 2 alleles combining X and Y for males. We filtered variants based on outputs from a support vector machine (SVM) classifier based on inferred pedigree of related and duplicated individuals to calculate Mendelian consistency statistics and other features^31,40^.

### Initial exclusions of individuals from analysis

#### InPSYght

We excluded individuals with: sex mismatches (self-reported sex disagreed with genetic sex) (n=20), non-XX or XY sex karyotypes (n=17), estimated DNA contamination >5% using verifyBamID2^41^ (n=4), or for whom <98% of sites were at a sequencing depth of ≥10 (n=14). See **Table S4**.

#### TOPMed

We excluded individuals with: sex mismatches, non-XX or XY sex karyotypes, or that had estimated DNA contamination > 5%^31^.

### Construction of principal components for sample inclusion and analysis

To compute principal components (PCs) of genetic ancestry, we removed variants in high long-range LD regions^42^ and then pruned variants in PLINK/1.9^43^ using --indep-pairwise flag and the following parameters: 500000 5 0.2. We computed PCs using the pca flag within PLINK^43^ and restricted variant selection to those present in the Human Genome Diversity Project (HGDP) reference panel^44^. These criteria resulted in 160,517 common (MAF>5% in the InPSYght case/control + TOPMed control dataset) autosomal bi-allelic SNVs (from the WGS data). We first retained InPSYght individuals that visually clustered in the first three PCs. Then, we retained TOPMed controls that visually clustered in the space occupied by the InPSYght individuals along the first three PCs (**Figure S1 A-C**).

### Identification of duplicated and related individuals

#### InPSYght and TOPMed

We estimated pairwise kinship among all individuals (InPSYght and TOPMed controls) using KING^45^. We excluded one person per pair of monozygotic/duplicate genotyped individuals (17 duplicates within InPSYght, 140 duplicates within TOPMed, and 52 within InPSYght/TOPMed). For InPSYght/TOPMed pairs, the InPSYght sample was retained. To obtain unrelated individuals for variant aggregation tests we randomly retained an individual from related pairs or groups (defined by individuals being related with at least one other person in the group) with kinship > 0.0409 (third degree relationship or higher) resulting in 4489 cases (3006 SZ and 1483 BD), 2374 InPSYght controls, and 8509 TOPMed controls.

### Genetic ancestry estimation in InPSYght using WGS data

We subsequently estimated global genetic ancestries for InPSYght samples using WGS data. We used the supervised learning approach implemented in the software ADMIXTURE^46^ trained on the 1000 Genomes Project phase 3^47^ ancestry super-populations: Admixed Americans (AMR), African (AFR), European (EUR), South Asian (SAS), and East Asian (EAS). To estimate sample ancestry, we used the same SNV’s as for the PC analysis.

We further inferred chromosomal-level ancestry using individuals of European and African origins from the HGDP reference panel. HGDP was chosen here in an attempt to characterize known high-levels of genetic diversity across African populations^48^. From the HGDP reference, we used 156 European individuals as a single group and as separate groups we used between 8 and 27 individuals per population from the following eight African populations: Bantu from South Africa, Bantu from Kenya, Biaka Pygmy, Mandenka, Mbuti Pygmy, Mozabite, San, and Yoruba^49^.

### Genetic ancestry estimation in TOPMed using WGS data

Estimated genetic ancestry of TOPMed samples has been previously described^31^. In summary, first, local ancestry was inferred using RFMix v2^50^ with the following option: --node-size=5. For reference haplotypes used in local ancestry inference, we obtained the Human Genome Diversity Panel (HGDP)^51^ and processed the data according to Wang et al. (2014)^52^, end up with 938 individuals and 639,958 autosomal SNVs. We then condensed the 53 populations in HGDP into 7 super-populations: (1) Sub-Saharan Africa (n=104), (2) Central/South Asia (n=200), (3) East Asia (n=229), (4) Europe (n=154), (5) Native America (n=63), (6) Oceania (n=28), (7) Middle East (n=160). After running RFMix, we summed up inferred local ancestry across all genetic windows of each individual to calculate global ancestry proportions, corresponding to the seven super-populations. Almost all selected TOPMed controls (n = 14,804, 99.9%) had >25% African estimated global ancestry (range: 26% to 100%) except 8 (0.05%) selected controls; range of estimated African ancestry: 8% to 24%).

### Power calculations to detect single-variant associations

We calculated the odds ratio (OR) that would yield 80% power to detect variants associated with BD, SZ, or SZ + BD, using InPSYght + TOPMed samples as controls. We assumed a disease prevalence of 1% for BD and 1% for SZ and 2% for SZ + BD. We conservatively used the estimates of the number of unrelated cases (n=1500 BD and n=3000 SZ) and controls (n=11,000) present in the case/control comparison group of interest in our calculations. We assumed risk alleles frequencies of 0.01 and 0.05, a multiplicative disease model on the OR scale, population-based controls, and genome-wide significance level of 5×10^−9^ ^53^.

### Genome-wide single-variant case-control association analysis

We tested for association of SZ and/or BD with each SNV or indel (minor allele count (MAC) >20 in the tested individuals) using SAIGE (version 0.42), which employs a mixed model to account for related individuals and uses saddlepoint approximation to account for case-control imbalance in estimation of significance^54^. We used the reference genome allele as the reference allele in calculation of Odds Ratios (OR). We performed a total of six case-control association analyses (Figure 1). We used as cases the SZ only, BD only, or combined SZ or BD InPSYght samples. We used as controls either only the InPSYght controls or the InPSYght + TOPMed controls. We chose to combine the SZ and BD cases based on previous evidence of substantial genetic correlation between the two disorders (based on common variants in European-ancestry populations)^55^. We included as covariates genetic sex and the first ten genetic PCs. Using only the selected TOPMed and InPSYght individuals, we made 10 genetic PCs for analysis (as described in the section above) (**Figure S1 D-F**). We also included the sequencing batch as a covariate for InPSYght sample only analyses. To assess potential differences between the two sets of controls (InPSYght and TOPMed controls), we performed association analysis of InPSYght controls versus TOPMed controls. We controlled for multiple testing within an analysis group using p < 5 × 10^−9^ for genome-wide significance; we used p < 5 × 10^−9^ / 7 comparisons = 7.1 × 10^−10^ for a conservative genome-wide significance.

### Protein Truncating Variants Singletons-Based Burden Tests

We performed burden tests for protein truncating SNV or indels (PTV) singletons at both the gene level and gene-set level. We restricted our analysis to KING-estimated unrelated samples (less than third degree relationships, see above) consisting of 4489 InPSYght BD and SZ cases, 2374 InPSYght controls and 8509 TOPMed controls. We defined singleton variants based on the unrelated samples to avoid excluding variants that occurred multiple times in a single family. We annotated 61,732 singleton variants as PTV using the following Ensembl Variant Effect Predictor categories: frameshift, stop gained, splice acceptor, or splice donor. We performed gene-level burden tests of SZ + BD cases versus InPSYght + TOPMed controls on the aggregated singleton PTVs within each gene, testing for association of PTV count with case status using RVTESTS^56^. To maximize power, we restricted testing to the genes with > 10 PTV singletons (1045 of 22,178 genes) and only tested SZ + BD cases versus InPSYght + TOPMed controls given the limited number of PTVs available per gene for testing. We included as covariates the first ten genetic PCs, sex, and an individual’s total number of singleton alleles. We applied a Bonferroni-corrected significance threshold of 0.05/1045 [of genes with >= 10 PTV singletons]= 4.8 ×10^−5^.

In addition, among the 10 previously reported SZ associated genes from the SCHEMA consortium study^16^, we tested the 6 genes with at least one PTV singleton in our study; all six genes had < 10 PTV singletons. We note that the SCHEMA consortium study includes the InPSYght SZ and control samples; thus, this is not an independent test, but a test to see the contribution of the African American samples.

We performed gene set level tests for the enrichment of PTV singletons within sets of genes previously associated with SZ. Given the higher singleton counts in gene-set tests compared to individual genes, in addition to testing SZ + BD versus InPSYght + TOPMed controls as for individual genes, we also tested SZ versus InPSYght + TOPMed controls. We tested three gene sets previously associated with SZ in more than one study. Three gene sets were associated with SZ in more than one study: 1423 postsynaptic density genes^57,58^ and 784 FMR1 protein associated (formerly named FMRP) genes^59,60^ and 3063 constrained (pLI > 0.90) genes^60^. In addition, we tested a gene set we constructed containing the ten SCHEMA SZ associated genes^16^, which as noted above has some overlapping samples with the samples for this study. For each gene-set test, for each person we summed the PTV counts over all genes in the gene set and used RVTESTS^56^ to test for association with case status as described for single gene tests. We used a single analysis Bonnferoni significance threshold of 0.05/3=0.017; in analysis where a result passed the single analysis threshold we further evaluated the result using more stringent threshold 0.05/(3*2 comparisons)=0.0085 to account for multiple analyses.

### Construction of sequencing metadata principal components (metadata PCs)

To control for potential sequencing batch effects within and across InPSYght and TOPMed, we constructed PCs based on a shared set of sample-level sequencing quality control metrics, including per-sample average depths and sample contamination levels (**Table S5**). We used the first four of these sequencing metadata PCs in the chromatin and conservation states analysis.

### Test for case-control and InPSYght control-TOPMed control enrichment of rare and low-frequency variants in chromatin states and conservation states

We tested if cases and controls exhibit differential enrichment of rare and low-frequency SNVs (MAF < 0.05) for any class of genomic region defined based on chromatin or conservation states. Specifically, for the chromatin states, we used the universal ChromHMM^61^ 100-chromatin state annotation of the human genome, which captures combinatorial and spatial patterns of chromatin marks over 1000 epigenomic datasets from more than 100 cell and tissue types. The version of the annotations we used had been previously lifted over to human hg38 assembly from hg19^61^. For the conservation states, we used a ConsHMM^62^ 100-conservation state annotation of the human genome defined directly in hg38, which captures combinatorial and spatial patterns of individual nucleotides aligning to and matching the human reference genome within a 100-way vertebrate sequence alignment^62,63^.

For the same set of unrelated samples as in the gene-based tests, we used SNVs with MAF < 0.05, excluding variants overlapping ENCODE excluded regions^64^. We annotated each variant with the ChromHMM and ConsHMM annotations described above. For each of the six case/control and InPSYght control/TOPMed control comparisons described in the single-variant test section, we used logistic regression to test for association between the non-reference allele count (predictor) and case/control or control study status (outcome). We upweighted rarer variants with the beta function *beta*(MAF, 1, 25) (mirroring the default choice of WGScan^65^). We included as covariates, the first ten genetic PCs, sex, sequencing batch (for tests involving only InPSYght samples), and the weighted total count of rare and low-frequency variants for each sample as covariates. We repeated the analysis including the first four sequencing metadata PCs as covariates. We controlled for multiple testing with a Bonferroni correction for 200 tested states with significance p-value threshold calculated as 0.05/200 =0.00025; we used a more stringent p-value threshold 0.05/(200*7 comparisons) =3.6 × 10^−5^ to account for multiple analyses.

### Test for InPSYght control-TOPMed control enrichment of rare and low-frequency variants in various genomic repeat categories

To investigate the potential effects of between InPSYght and TOPMed study sequencing technical differences on the number of non-reference alleles detected in genomic repeat regions, we annotated each analyzed variant for its presence in a repeat region. We defined repeat regions using (1) repeat regions identified by RepeatMasker 3.0.1 obtained from the UCSC genome browser^66^ and (2) simple repeats defined by Tandem Repeats Finder^67^. The repeat regions were tested as a class and were further divided into 21 repeat categories, for a total of 22 categories. We tested for differential enrichment of SNVs in each of the repeat categories, without inclusion of the sequencing metadata PCs as described for the ChromHMM and ConsHMM state tests. We controlled for multiple testing with a Bonferroni correction for 22 tested repeat categories with significance p-value threshold calculated as 0.05/22=0.0023; we used a more stringent p-value threshold of 0.05/(22*7 comparisons)= 3.3 × 10^−4^ to account for multiple analyses.

### Genome-wide rare and low-frequency and rare variant sliding window burden tests for case-control and InPSYght control-TOPMed control comparisons

To identify local enrichments of disease-associated rare and low-frequency alleles we performed sliding window burden tests using WGScan^65^ in the same six unrelated-samples case/control sets and one unrelated control/control sample set as in the chromatin and conservation states analysis. We also used the same variants as in the chromatin and conservation states analysis. Following the default parameters of WGScan, we tested variants in window sizes of 5, 10, 15, 20, 25, and 50 kb (including coding and non-coding regions), and upweighting rarer variants with beta function *beta*(MAF, 1, 25) weights. We included the same set of covariates as in the chromatin and conservation states analysis (with and without the first four sequencing metadata PCs). We used WGScan’s permutational approach with default parameters (including 5000 permutation replicates) to estimate the effective number of tests (*n*) for each comparison group^65^. We controlled for multiple testing with a Bonferroni-type correction with significance thresholds calculated as 0.05/*n* (2.15 ×10^−8^ to 2.19 × 10^−8^); we used a more stringent p-value threshold 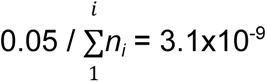 to account for multiple analyses sets (i), where 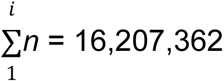 is the total number of effective tests across all comparisons.

### Secondary analysis for the most strongly associated window across all case-control test combinations

We conducted secondary analyses for the most strongly associated sliding window across all six case-control test combinations: the chr11:64,859,972-64,869,939 association observed in InPSYght BD versus InPSYght controls. We removed variants in the repeat regions (defined above) and performed a WGScan-based burden test of InPSYght BD versus InPSYght controls on this window using the approach described above. We also tested the InPSYght BD cases versus InPSYght controls single variant association in the non-repeat region of chr11:64,859,972-64,869,939 using two-sided Fisher’s exact test implemented in PLINK 1.9^43^ (as many variants had MAC lower than the SAIGE threshold (MAC<20)). We then performed two additional WGScan-based burden tests for this chr11:64,859,972-64,869,939 region: variants with nominally significant Fisher’s exact p-values (p-value < 0.05) only and variants in the window that were not nominally significant.

## Results

### Genome-wide single-variant case-control association analysis

The InPSYght study sample consists of 7544 African American individuals (estimated African ancestry > 25%): 1598 with BD, 3295 with SZ, and 2651 without known BD or SZ or unipolar depression (**Table S1**). 42% of the participants were female and participants had an average age of 42.5 +/− 12.7 years (**Table S3**). We generated WGS data for InPSYght samples at an average depth of 26.8 +/−5.5. We estimated the ancestral sources of African ancestry in the samples using the Human Genome Diversity Project (HGDP) reference panel and found that almost all InPSYght individuals were genetically most similar to the West African populations represented by Yoruba and Mandenka samples (**Figure S2**).

To increase power to detect BD- and SZ-variant associations, we included, as controls, 14,812 African American individuals from the TOPMed study (of which 99.9% had >25% African Ancestry (average sequencing depth 36.8+/−4.7). The TOPMed samples came from studies focused on disease of heart, lung, blood and sleep disorders and inclusion was not predicated on information about mental health (**Table S2**). To minimize differences between InPSYght and TOPMed samples used in our analyses, we jointly called the samples and we selected TOPMed samples to have a similar genetic PC composition as the InPSYght samples (**Figure S1, Methods**). The selected TOPMed samples were 62% female. In the jointly called InPSYght and TOPMed external control dataset, we identified 226,434,324 variants (210,210,658 SNVs and 16,223,666 short indels) on the autosomes and chromosome X, of which 220,310,579 variants have MAF < 0.05 (204,467,345 SNVs and 15,843,234 short indels).

To identify SNV and short indels associated with BD and SZ, we performed GWAS single-variant tests of association (MAC>20 in the tested group). We adjusted for the first 10 genetic PCs, sex, and sequencing batch (only in InPSYght sample analysis) and accounted for relatedness using a mixed model. First, to determine whether differences in genetic ancestry or sequencing between InPSYght and TOPMed samples might cause artifactual associations in the BD and SZ association analysis, we performed a GWAS of InPSYght controls versus TOPMed controls. There was no evidence of inflation of genomic control (λ_gc_=1.02). There was one common genome-wide significant variant, an indel on chromosome 13 (rs11350613) (OR (95%CI)= 0.79 (0.74, 0.85), p-value = 1.2 × 10^−10^, MAF of 0.63 versus 0.68 in InPSYght controls versus TOPMed controls, respectively) (**Figures S3**). However, variant rs11350613 is not in LD with other variants in our data, and it just barely passed the SVM-based QC filter in our study (−0.497, threshold for retention SVM > −0.5) and failed QC in the subsequent TOPMed 10 data freeze (https://bravo.sph.umich.edu/variant.html?chrom=13&pos=79615934&ref=C&alt=CT). Next, we estimated the power to detect case/control associations for each case group versus the InPSYght + TOPMed control group. We used p < 5 × 10^−9^ for genome-wide significance and p < 5 × 10^−9^ / 7 = 7.1 × 10^−10^ for a conservative multiple groups testing genome-wide significance to account for the seven groups of samples being tested. For tests of BD, SZ and SZ + BD association with InPSYght + TOPMed controls we have approximately 80% power for p <5 × 10^−9^ and p< 7.1 × 10^−10^ to detect ORs of 3.2, 2.4 and 2.1, and 3.4, 2.5 and 2.3 respectively, for minor allele frequency (MAF) of 0.01 and for ORs of 1.76, 1.53 and 1.43, and 1.78, 1.55 and 1.44, respectively for MAF of 0.05, respectively. We performed single-variant association analyses of BD, SZ or SZ + BD versus InPSYght + TOPMed controls or versus InPSYght controls (Manhattan and QQ plots **Table S6**, **Figures S3-S9**). Estimation of population stratification and deviation of test statistics observed from that expected (λ_GC_) ranged from 1.00 to 1.02 for the various case-control combinations tested, consistent with minimal stratification bias and p-value inflation (**Table 1**). We observed one genome-wide significant, but not multiple group testing corrected genome-wide significant (between p < 5 × 10^−9^ and p < 5 × 10^−9^ /7 comparisons = 7.1 × 10^−10^), locus on chromosome 18 (two SNVs and one indel) in the BD versus InPSYght + TOPMed control analysis (**Figures S6**): lead SNV chr18:49738979:G:T, OR (95%CI)=30.7 (1.35 × 10^−9^), p-value= 1.3 × 10^−9^, MAF of 0.0069 (BD) versus 0.0011 (InPSYght + TOPMed controls. The three variants are within 600 bp of each other and are in strong LD (r^2^ > 0.9) (Figure 2). The locus zoom plots display the 1000G AFR LD and there appears to be a variant in r^2^ > 0.80 with our associated variants; however in our data, this variant has r^2^=0.65 with the most strongly associated variants and p-value=0.0014. We observed less significant association results in the smaller BD versus InPSYght control analysis (chr18:49738979:G:T, OR (95% CI)=8.51 (3.85-18.8), p-value = 1.22 × 10^−7^, MAF of 0.0069 (BD) versus 0.00056 (InPSYght controls) (**Table S7**). These variants had no obvious QC issues. The nearest genes in the region are *ACAA2*, *LIPG*, and *MYO5B*.

**Figure 2.**
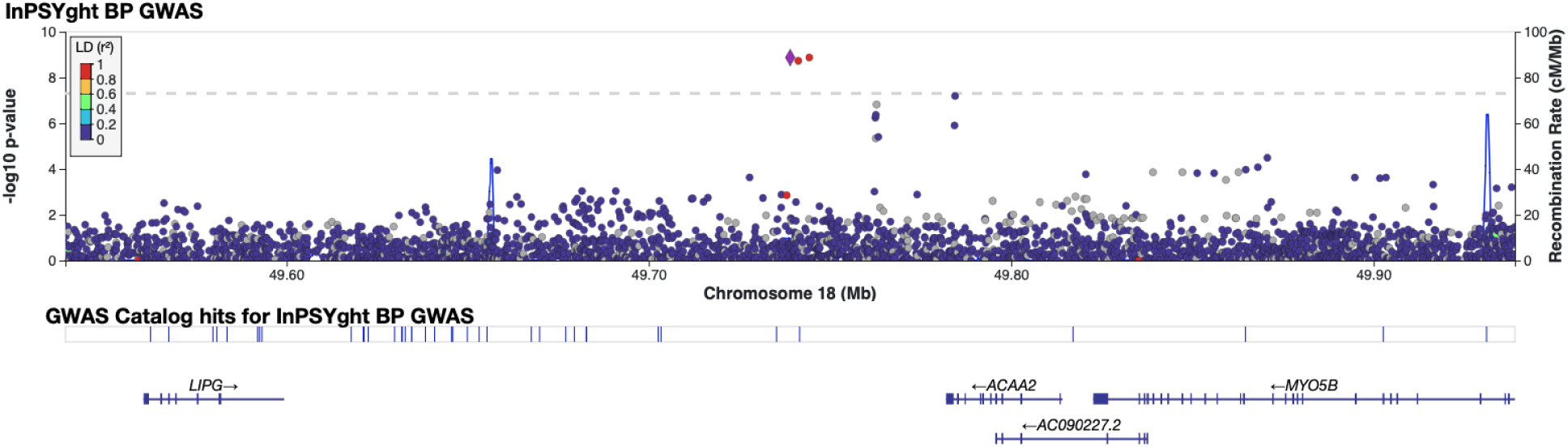
Regional view of chromosome 18 locus showing evidence of association in the InPSYght BD versus InPSYght+TOPMed controls. Horizontal line shows genome-wide significance threshold of 5 × 10^−9^.

**Table 1.** Lambda GC for each analysis type. (S) represents at least one genome-wide (p <5×10^−9^) association. *Comparison was not tested in this study. Base refers to the association test with base covariates (see Methods) without the inclusion of Metadata PCs. BD = Bipolar disorder; SZ = Schizophrenia

| Lambda GC results<br>((S) denotes a significant association) |  |  |  |  |  |  |  |
| --- | --- | --- | --- | --- | --- | --- | --- |
| Sample group 1 | Sample group 2 | Single variant GWAS | Gene exon burden | Chromatin/ Conservation burden |  | Sliding window burden |  |
|  |  | Base | Base | Base | + Metadata PCs | Base | + Metadata PCs |
| InPSYght controls | TOPMed controls | 1.02 (S) | * | 1.51 | 1.02 | 1.00 | 1.01 (S) |
| InPSYght BD + SZ | InPSYght + TOPMed controls | 1.01 | 0.92 | 1.54 | 1.24 | 1.02 | 1.01 |
|  | InPSYght controls | 1.00 | * | 0.98 | 0.98 | 0.98 | 0.98 |
| InPSYght BD | InPSYght + TOPMed controls | 1.02 (S) | * | 1.70 (S) | 1.21 | 1.00 | 0.99 |
|  | InPSYght controls | 1.00 | * | 1.13 | 1.07 | 0.98 | 0.98 |
| InPSYght SZ | InPSYght + TOPMed controls | 1.00 | * | 1.28 | 1.10 | 1.02 | 1.01 |
|  | InPSYght controls | 1.00 | * | 0.92 | 0.93 | 0.99 | 0.98 |

We found that in chromatin and conservation state tests (below), sequencing metric-based PCs may help control for sample sequencing differences. We repeated our analysis of this region including four sequencing metric metadata PCs. We found slightly attenuated non-genome-wide significant results (chr18:49738979:G:T, OR (95% CI)=15.0 (5.78-39.1), p-value 2.7 × 10^−8^).

### Gene based and gene set tests

The SCHEMA consortium exome-sequencing meta-analysis of 24,248 SZ cases and 97,322 controls of predominantly European ancestry individuals tested functional variant annotation groupings for association with SZ; they found extremely rare PTVs had the strongest SZ associations^16^. The SCHEMA analysis contained InPSYght SZ and control individuals, but did not separately report the results for InPSYght African American individuals. To specifically assess gene-based PTV associations in African American individuals, we used 4489 InPSYght SZ + BD cases and 10,883 InPSYght + TOPMed controls (all unrelated individuals) to test for PTV burden in 1045 genes with singleton PTV count > 10 (see **Methods**). We included the BD cases to increase the number of alleles per gene given similarities in the underlying genetic architecture^55^. We did not detect evidence of inflation of association statistics (λ_GC_ = 0.92) and found no SZ+BD associated genes (**Table 1, Figure S10**). Of SCHEMA’s ten most strongly associated SZ genes: four genes had no singleton PTVs in the InPSYght + TOPMed sample and six had singleton PTV <10; all had p>0.05 (**Table S8**). Considering SCHEMA’s top ten genes as a single gene set, we observed directionally consistent, though non-significant, enrichments of PTVs in SZ (OR=1.65, p=0.42) and SZ + BD cases (OR=2.07, p=0.15) compared to controls.

We tested three previously identified SZ-associated gene sets for PTV gene set enrichment in SZ or SZ+BD versus InPSYght + TOPMed controls. For the most strongly enriched SCHEMA^16^ gene set - 3063 constrained (pLI > 0.90) genes^16,60^ - we found significant association for both SZ (OR=1.11, p=8.2×10^−3^) and SZ + BD (OR=1.13, p=2.9×10^−3^) (samples contained within SCHEMA). For two SZ-associated gene sets identified in multiple papers - 1423 post-synaptic density genes^57,58^ and 784 FMR1 protein associated (formerly named FMRP) genes^59,60^ - we found directionally consistent OR’s but no significant associations (**Table 2**).

**Table 2.** InPSYght singleton PTV burden test results for previously published schizophrenia gene sets. ^#^Overlap in study sample with InPSYght; *Gene set was not tested in this study.

|  |  | Study<br>(Disorder) | InPSYght<br>(BD&SZ) | InPSYght<br>(SZ) | Genovese<br>et al<sup>58</sup><br>(SZ) | SCHEMA<sup>16</sup><br>(SZ) |
| --- | --- | --- | --- | --- | --- | --- |
|  |  | Case N<br>Control N | 4489<br>10,883 | 3006<br>10,883 | 4877<br>6203 | 24,248<br>97,322 |
|  |  | Ancestry | African<br>American | African<br>American | European | Multi-ethnic |
| Gene Set | #<br>Genes |  | OR<br>(P value) | OR<br>(P value) | OR<br>(P value) | OR<br>(P value) |
| Constrained Genes<br>(pLI > 0.90) | 3063 |  | 1.13<br>(0.0030) | 1.13<br>(0.0082) | 1.17<br>(1.7x 10<sup>-8</sup>) | 1.26<br>(7.6x 10<sup>-35</sup>) |
| PSD Genes | 1423 |  | 1.09<br>(0.052) | 1.11<br>(0.039) | * | 1.20<br>(1.0x10<sup>-6</sup>) |
| FMRP<br>Genes | 784 |  | 1.10<br>(0.058) | 1.10<br>(0.099) | 1.23<br>(8.2x 10<sup>-9</sup>) | 1.25<br>(2.2x 10<sup>-17</sup>) |

### Case-control enrichment of rare and low-frequency SNVs in chromatin states and conservation states

We next investigated whether chromatin or conservation state-based sets of rare and low-frequency SNVs (MAF < 0.05) are differentially enriched between the InPSYght and TOPMed controls, and between SZ and/or BD cases and InPSYght and/or TOPMed controls (Figure 1, unrelated individuals). Our goal with this analysis was to test a diverse set of systematically defined genomic regions that could potentially capture both technical artifacts or biological associations. For this we annotated the SNVs using a set of 100-ChromHMM universal chromatin states^61^ and a set of 100-ConsHMM conservation states^62,63^, for a total of 200 states (Methods). For each state we performed a logistic regression to test if two groups have a differential variant burden, with upweighting of variants with lower MAF (**Methods**).

We first tested for state differences in variant burden in InPSYght controls versus TOPMed controls and found that the p-value distribution was substantially inflated compared to the expected distribution (lambda GC =1.51, Figures 3AB, **Table 1**). Likewise, in case-control analyses that included TOPMed controls, we found substantial p-value inflation (lambda GC =1.28-1.70, **Table 1**, Figures 3CD**, Figure S11**). One state (ConsHMM state 75) had lower variant burden in BD cases compared to InPSYght + TOPMed controls (OR=0.71, p=9.9×10^−5^, significant at a per case-control group level).

**Figure 3.**
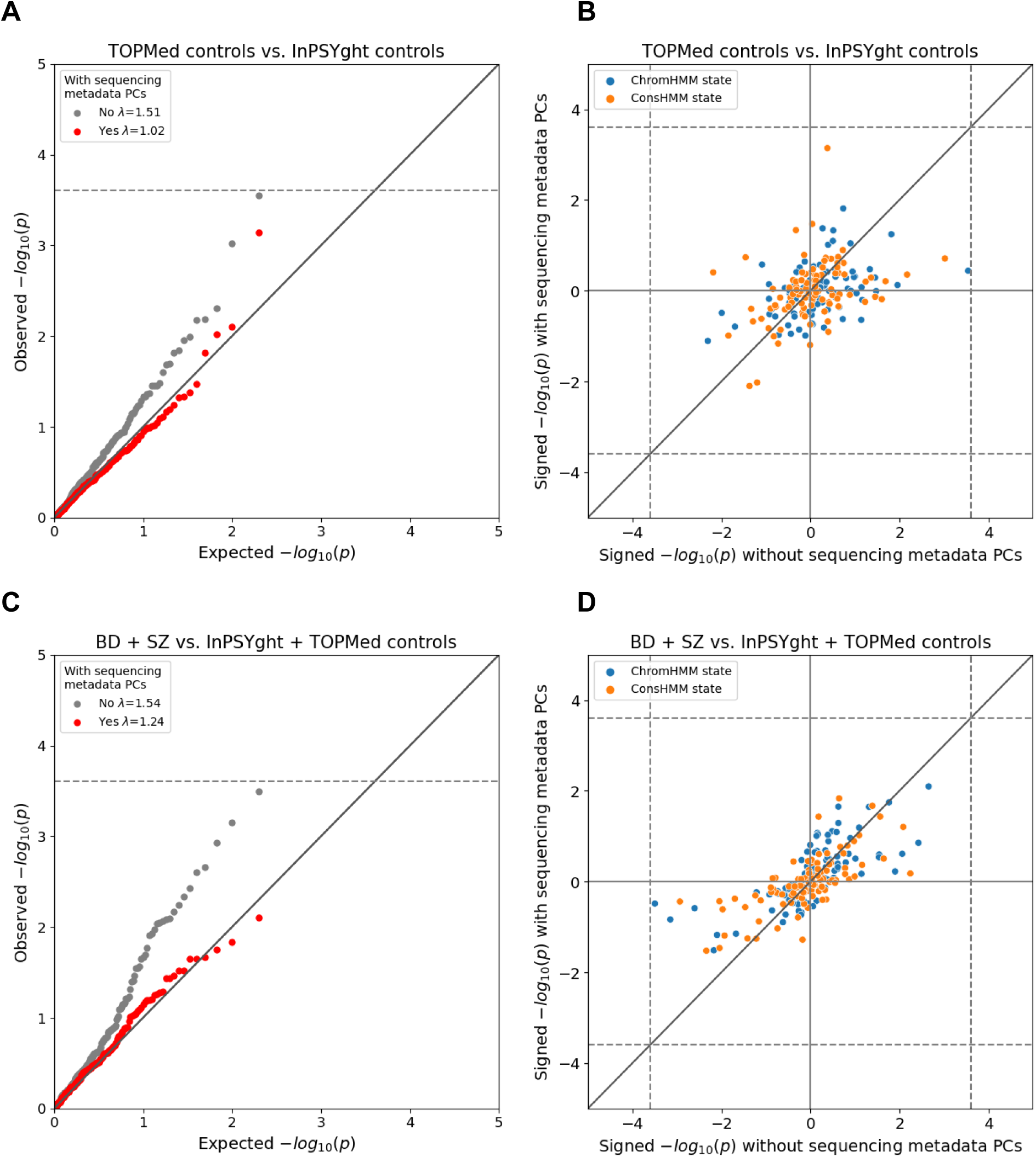
Chromatin and conservation state burden test results for InPSYght cases versus InPSYght controls (A, B) and InPSYght cases versus InPSYght and TOPMed controls (C, D), with each point representing a ChromHMM or a ConsHMM state. Dashed lines show Bonferroni-based p-value thresholds (p=0.05/200). Diagonal lines show the unit slope. (A, C) QQ-plots with genomic inflation factors (λ) before and after inclusion of sequencing metadata PCs covariates. (B,D) Signed (by direction of enrichment coefficient) -log10 p-values before and after inclusion of sequencing metadata PCs covariates. Sign direction: Case enrichment values are positive, control enrichment values are negative.

To test whether these findings could be due to differences in technical sequencing factors between InPSYght and TOPMed we constructed a set of sequencing metadata PCs from sequencing quality control metrics (**Methods**) and included them as covariates. These sequencing metadata PCs summarize various quality control metrics, including various sequencing depth-related metrics that almost exclusively drove the PCs (**Table S5**). After inclusion of sequencing metadata PCs, the TOPMed control versus InPSYght controls state-based burden test had a non-inflated lambda GC =1.02 and the TOPMed control-containing case/control analyses had lower lambda GCs (1.10-1.24) with no visible inflation at more significant p-values (**Table 1**, Figure 3BD**, Figure S11**). Across all the case-control comparisons no state reached a Bonferroni-based significance threshold of 0.05/200 chromatin states = 2.5 × 10^−4^ (the number of total states tested between the two models) after the inclusion of metadata PCs (**Figure S12**). Thus, we did not find evidence for enrichment in BD and/or SZ cases of rare and low-frequency variants in particular chromatin or conservation states.

### Control-control enrichment of rare and low-frequency variants for repeat classes

We next sought to better understand why the inflated lambda GC values for chromatin and conservation states in the control-control enrichment analysis had non-inflated lambda GC when including sequencing metadata PCs. Given repeat regions often present technical challenges in sequencing, we hypothesized that repeat regions as a whole or of particular classes might show specific differences in enrichment by control study. This hypothesis would also be consistent with strong enrichments shown for various categories of genomic repeat elements in specific ChromHMM^61^ and ConsHMM^62^ states. To test this hypothesis, we annotated rare and low-frequency SNVs with a total of 21 different categories of repeat regions defined by RepeatMasker and Tandem Repeats Finder^67^. Using the same test as for state-based analysis, we asked if the TOPMed and InPSYght controls were differentially enriched for SNV over all repeats and in specific repeat categories. We did not see significant enrichment of SNV in the overall repeat category (OR=2.13, p=4.1 × 10^−2^, overlapping 53% of variants on average). However, we found significant enrichment of rare and low-frequency variants in TOPMed controls compared to InPSYght controls in the RepeatMasker-defined SINE (OR=2.04, p=3.0 × 10^−9^, overlapping 16% of variants on average) and simple repeat regions (OR=1.31, p=3.2 × 10^−6^, overlapping 1% of variants on average) (Figure 4AB). After including sequencing metadata PCs as additional covariates, only simple repeat regions remain significant (at a single analysis set level), with a greatly reduced significance level (OR=1.35, p=2.0 × 10^−3^) (Figure 4CD). Overall, these results suggest that TOPMed and InPSYght cohorts have different distributions of variants in specific repeat categories and could be associated with artifactual associations in burden-type analyses. Such difference can be partially controlled for with the inclusion of sequencing metadata PCs.

**Figure 4.**
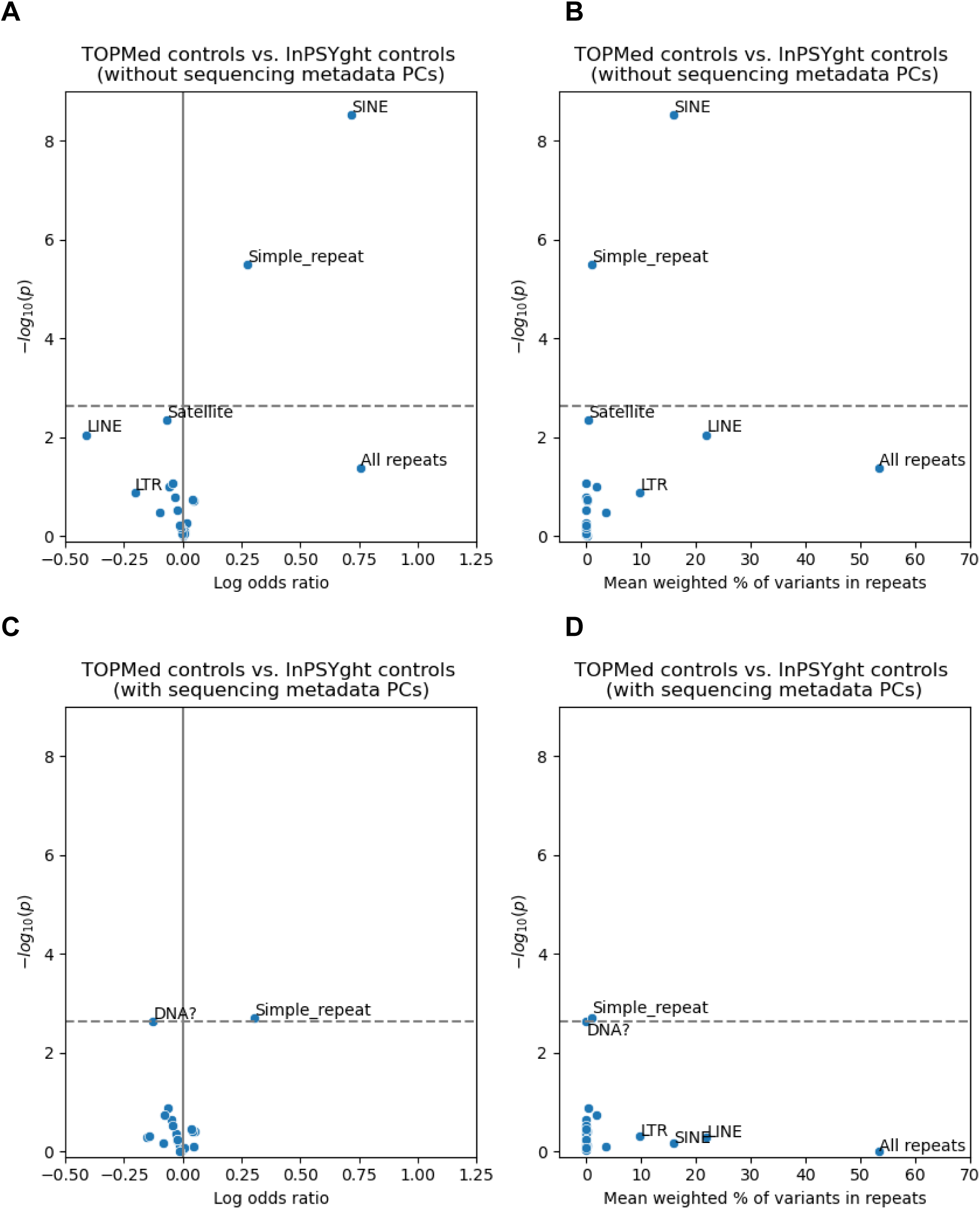
Test of repeat categories for enrichment of rare and low-frequency variants in TOPMed controls versus InPSYght controls, without (A, B) or with (C, D) sequencing metadata PCs covariates. “DNA?” represents elements with uncertain category classification to the DNA repeat element category. Horizontal dashed lines show Bonferroni-based p-value thresholds (p = 0.05/22). (A), (C): volcano plots with log odds ratio on the x-axis and -log10 p-values on the y-axis. (B), (D): Mean % of variants overlapping each repeat category on the x-axis (weighted by the minor allele frequencies, Methods), and -log10 p-values on the y-axis.

### Genome-wide rare and low-frequency variant sliding window burden tests for BD and SZ samples versus controls

To identify contiguous genomic regions that might harbor an excess of rare and low-frequency SNVs that predispose to or protect from BD and SZ, we conducted genome-wide 5-50kb sliding window analyses within the TOPMed and InPSYght control groups and six case-control groups (Figure 1) using the WGScan framework^65^. Within each genomic window we performed allele frequency weighted burden test, adjusting for covariates used in the chromatin and conservation state analysis (with and without the metadata PCs).

To identify windows that might be affected by differences in sequencing between the TOPMed and InPSYght study we first compared InPSYght controls and TOPMed controls (Figure 5). The lambda GC =1.01 was consistent with no inflation of the test statistics (**Table 1**). However, we identified a 50kb region on chromosome 5 (chr5:58,500,010 - 58,550,007) with 12 windows with genome-wide significant associations with the strongest association being chr5:58,510,332 - 58,515,331 (p=1.29 × 10^−9^, significant when accounting for multiple analysis sets (Figure 5B). This region showed an elevated burden of rare and low-frequency SNVs for InPSYght controls (mean weighted burden=156) compared to TOPMed controls (mean weighted burden=145). To test the robustness of this association we repeated the analysis without including the sequencing metadata PCs as covariates, and found that none of the windows in this region remained significant (minimum p=2.16 × 10^−3^) (Figure 5ACD), leaving open the question of if this was a chance association or if the metadata PC’s induced a false positive association in this region.

**Figure 5.**
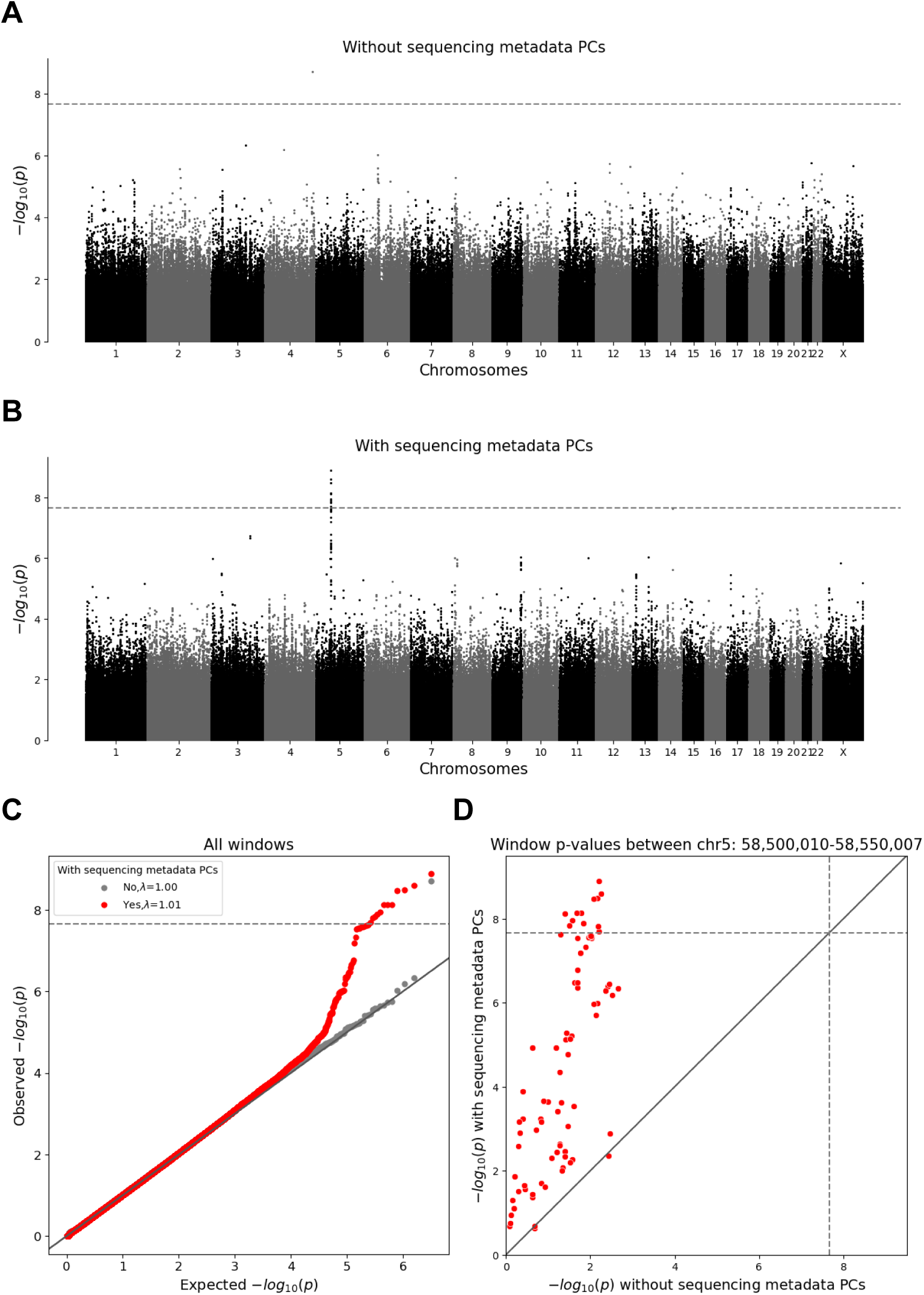
WGScan genome-wide sliding window burden test for rare and low-frequency variants for InPSYght controls versus TOPMed controls. Horizontal lines show genome-wide p-value significance thresholds (p=2.18 × 10^−8^). Diagonal line shows the unit slope. (A, B) Manhattan plots of sliding window p-values without sequencing metadata PCs (A) and with sequencing metadata PCs (B). (C) QQ-plot for the window p-values, before and after inclusion of sequencing metadata PCs covariates. (D) Comparison between sliding window p-values before and after inclusion of sequencing metadata PCs covariates, showing windows in the chr5 region 58,500,010-58,550,007 (red).

In the six case-control analyses we observed no inflation of the test statistics (lambda GC = (0.98-1.01)) with the inclusion of metadata PCs (**Table 1**). We did not identify any window where the burden of rare and low-frequency SNVs was significantly associated with BD and/or SZ. The significant chromosome 5 region from the control-control analysis did not show association signal in any of the case-control analyses (p>=0.01 for every case-control analysis).

Interestingly, when we compared the sliding window results to BD and SZ GWAS results, we noticed that the most significant window across all case-control tests was located in a 6Mb region of chromosome 11 containing multiple independent PGC common variant BD associations (**Table S9**)^4^. This 10kb window overlaps the *EHD1 gene* (chr11:64,859,972-64,869,939; GRCh38 (p=4.06 × 10^−8^ for InPSYght BD versus InPSYght controls and has a higher burden in controls than in cases). This window is the tenth most significant window for the InPSYght BD versus combined InPSYght and TOPMed controls analysis (p=1.37 × 10^−6^).

We assessed whether removing non-repetitive regions might strengthen the association signal because we could more accurately genotype non-repeat variants. In the non-repetitive region analysis of InPSYght BD versus InPSYght controls including sequencing metadata PCs, this window reached genome-wide significance (p=1.12 × 10^−9^) and was the most strongly associated window across all case-control comparisons; we found similar results when the analysis was run without metadata PCs (p=1.09 × 10^−9^). We identified a specific subset of control-enriched variants belonging to distinct haplotypes (**Figure S13**). Across cell and tissue types, this 10kb window has an average of 44.5% of basepairs in the TxReg chromatin state (defined by a high presence of transcription, enhancer, and promoter chromatin marks) from a chromatin state model providing per-cell and tissue annotations^68^; only 0.02% of 10kb windows in the genome had a higher percent of basepairs annotated by this state suggesting high regulatory potential (**Figure S14**; see **Supplemental Text** for further analysis). These analyses suggest a potential convergence of BD-related associations in this region that awaits replication in larger samples.

## Discussion

We investigated the genomic basis of SZ and BD across the allele frequency spectrum, for coding and non-coding variants in a cohort of African American individuals. We identified three low-frequency BD-associated variants (two SNVs and one indel) on chromosome 18. These variants are 200 kb from a BD association identified in a GWAS study of 1461 BD cases (Bipolar 1 disorder) and 2008 controls^38^; larger studies have not identified associations in this region^4,12^. In addition to the single-variant results, in a secondary BD analysis excluding genomic repeat regions we identified a significantly associated window on chromosome 11. The associated window is located in a 6 MB cluster of five common variant GWAS associations for BD^4^. This region has a significantly lower burden of rare and low-frequency and rare variants in BD cases compared to InPSYght controls. Given the number of GWAS and sliding window tests we performed (albeit with overlapping sets of cases and controls) these findings would likely not survive a more stringent correction for the effective number of tests performed in our study. Thus, we consider these chromosome 18 and 11 results as potentially associated loci that would need to be assessed in larger WGS studies including those with African American individuals.

Although we worked to minimize genotype calling differences by calling InPSYght and TOPMed samples together, one potential limitation of our study is that the TOPMed controls were more deeply sequenced and were sequenced separately from InPSYght cases and controls. Across our primary analyses, the only tests that appeared to be sensitive to potential differences in sequencing were the ConsHMM conservation states and of universal ChromHMM chromatin states tests (based on an inflated lambda GC). These tests combine large numbers of rare and low-frequency variants from regions that may be more or less difficult to sequence or prone to sequencing artifacts. These states are known to have different levels of repetitive regions^61,62^, and we found that particular classes of repeats, SINE and simple repeat region, were enriched for significant differences between the InPSYght and TOPMed controls. These findings suggest that sensitivity analyses should be performed for tests that aggregate variants across any class of genomic regions. For example, gene set analysis of non-coding variants may be susceptible to false positives given that different repeat classes are enriched near biologically distinct sets of genes; SINE are enriched in housekeeping genes^69^.

In our initial analyses of genomic regions, we attempted to control for sequencing differences and variant density, in a manner analogous to that of tests of rare singletons, by inclusion of the total weighted rare and low-frequency allele count as a covariate. This covariate was not effective in controlling the false positive rate in the ConsHMM conservation and ChromHMM chromatin states tests. We found, however, that inclusion of sequencing metadata PCs, reduced or even eliminated the lambda GC inflation. This suggests that for tests that aggregate rare and low-frequency variants (and potentially tests that aggregate singletons), inclusion of more extensive sequence metrics may be necessary to control the false positive rate.

In contrast to their ability to control for p-value inflation in the InPSYght versus TOPMed control chromatin and conservation state tests, we observed that the inclusion of the sequencing metadata PCs appeared to induce a false positive association in the sliding window control-control (TOPMed versus InPSYght) analysis. When we included sequencing metadata PCs as covariates, we identified a genome-wide significant region on chromosome 5 that was 6 orders of magnitude more significant than without sequence data metadata PCs. These findings suggest that significant associations should be subject to sensitivity analyses with and without adjustment for sequencing-related covariates. The findings also highlight the challenges associated with controlling for batch/sequencing effect differences and variant quality in WGS studies^70^.

There are additional limitations in our current study and analysis. First, we have a very small effective sample size relative to genotype array-based GWAS studies. We expect larger WGS case-control samples will be essential to identification of rarer non-coding variants.

Second, the TOPMed controls were not screened for psychiatric disorders, and thus the presence of individuals with BD or SZ in the control group could have decreased our power to detect BD and SZ associations. We expect misclassification in the control group to have a minimal effect^71^ because 1) the combined prevalence of BD and SZ is 2-4% in the general population^72^, and 2) individuals with severe mental illness are less likely to participate than individuals without severe mental illness in non-psychiatric disorder-based studies due to exclusion criteria and/or a decreased likelihood of enrollment^73,74^. As improved methods for analyzing rare non-coding variation are developed^65,71^, and the field comes to a clearer consensus on which analytical strategies and annotations are most effective for WGS analysis, more disease relevant information will be extracted from WGS data.

In summary, WGS allowed us to test variants for association with SZ and/or BD across the allele frequency spectrum in both the coding and non-coding regions, in particular rare non-coding variants which are missed by GWAS or exome sequencing studies. Our study highlights the need to perform sensitivity analysis when conducting WGS analyses, particularly for aggregation tests of rare and low-frequency non-coding variants that use data from multiple studies, and the need for larger WGS studies of African American individuals. We expect our data and additional African American WGS studies will contribute to the growing understanding of rare non-coding variants and complex psychiatric diseases in individuals with different ancestries.

## Data Availability

Whole genome sequence data for the GPC samples are available in ANVIL: WGSPD Project 1: Whole Genome Sequencing for Schizophrenia and Bipolar Disorder (WGSPD1). The Anvil Terra Workspace containing the data (https://anvilproject.org/data/studies/phs002041/workspaces) is AnVIL_NIMH_Broad_WGSPD1_McCarroll_Pato_GRU_WGS. Phenotype data is available at dbGaP Study Accession: phs002041.v2.p1.
Code for ChromHMM and ConsHMM state-based analysis are available at https://github.com/ernstlab/inpsyght_states

https://anvilproject.org/data/studies/phs002041/workspaces

## Acknowledgements

See Supplemental Acknowledgements for additional acknowledgements.

This work was supported by: University of Michigan Precision Health Scholar Award (https://precisionhealth.umich.edu/) (S.A.G.T.); R01 HG009976 (M.B.); U01MH105653 (M.B., C.N.P., S.A.M.); salary support from the Fonds de Recherche du Québec – Santé: a Junior 1 Award and currently a Junior 2 Award (S.A.G.T.); R01 MH115676 (R.A.O.) and (DP1DA044371 and U01HG012079) (J.E.)

This work used computational and storage services associated with the Hoffman2 Cluster which is operated by the UCLA Office of Advanced Research Computing’s Research Technology Group.

Bio-samples and data for the InPSYght study were obtained from NIMH Repository & Genomics Resource, a centralized national biorepository for genetic studies of psychiatric disorders. Contributing studies include the Genomic Psychiatry Cohort, the Consortium of the Genetics of Schizophrenia, the Biopolar Genome Study, the Lithium treatment moderate dose use study, and the Systematic Treatment Enhancement Program for Bipolar Disorder. We gratefully acknowledge the participants who provided biological samples and data for these studies.

We acknowledge the TOPMed Neurocognitive Working Group (members listed in the supplement).

Molecular data for the Trans-Omics in Precision Medicine (TOPMed) program was supported by the National Heart, Lung and Blood Institute (NHLBI).

Genome Sequencing for “NHLBI TOPMed: Cleveland Family Study - WGS Collaboration” (phs000954) was performed at NWGC (3R01HL098433-05S1).

Genome Sequencing for “NHLBI TOPMed: New Approaches for Empowering Studies of Asthma in Populations of African Descent - Barbados Asthma Genetics Study (BAGS)” (phs001143) was performed at Illumina (3R01HL104608-04S1).

Genome Sequencing for “NHLBI TOPMed: Genetic Epidemiology of COPD Study” (phs000951) was performed at NWGC and Broad Genomics (3R01HL089856-08S1, HHSN268201500014C, HHSN268201500014C).

Genome Sequencing for “NHLBI TOPMed: Multi-Ethnic Study of Atherosclerosis (MESA and MESA AA_CAC) “(phs001416.v1.p1) was performed at the Broad Institute of MIT and Harvard (3U54HG003067-13S1).

Genome Sequencing for “NHLBI TOPMed: Atherosclerosis Risk in Communities Study VTE cohort” (phs001211) was performed at Baylor (3U54HG003273-12S2 / HHSN268201500015C).

Genome Sequencing for “NHLBI TOPMed: Women’s Health Initiative” (phs001237) was performed at Broad Genomics (HHSN268201500014C).

Genome Sequencing for “NHLBI TOPMed: Hypertension Genetic Epidemiology Network” (phs001293) was performed at NWGC (3R01HL055673-18S1).

Genome Sequencing for “NHLBI TOPMed: Sarcoidosis in African Americans” (phs001207) was performed at Baylor (3R01HL113326-04S1).

Genome Sequencing for “NHLBI TOPMed: Mount Sinai BioMe Biobank” (phs001644) was performed at Baylor (HHSN268201600033I).

Genome Sequencing for “NHLBI TOPMed: Coronary Artery Risk Development in Young Adults” (phs001612) was performed at Baylor (HHSN268201600033I).

Core support including centralized genomic read mapping and genotype calling, along with variant quality metrics and filtering were provided by the TOPMed Informatics

Research Center (3R01HL-117626-02S1; contract HHSN268201800002I). Core support including phenotype harmonization, data management, sample-identity QC, and general program coordination were provided by the TOPMed Data Coordinating Center (R01HL-120393; U01HL-120393; contract HHSN268201800001I). We gratefully acknowledge the studies and participants who provided biological samples and data for TOPMed.

## Data and Code Availability

Whole genome sequence data for the GPC samples are available in ANVIL: WGSPD Project 1: Whole Genome Sequencing for Schizophrenia and Bipolar Disorder (WGSPD1). The Anvil Terra Workspace containing the data (https://anvilproject.org/data/studies/phs002041/workspaces) is AnVIL_NIMH_Broad_WGSPD1_McCarroll_Pato_GRU_WGS. Phenotype data is available at dbGaP Study Accession: phs002041.v2.p1.

Code for ChromHMM and ConsHMM state-based analysis are available at https://github.com/ernstlab/inpsyght_states

## References

1. Plana-Ripoll, O., Pedersen, C.B., Agerbo, E., Holtz, Y., Erlangsen, A., Canudas-Romo, V., Andersen, P.K., Charlson, F.J., Christensen, M.K., Erskine, H.E., et al. (2019). A comprehensive analysis of mortality-related health metrics associated with mental disorders: a nationwide, register-based cohort study. The Lancet 394, 1827–1835. 10.1016/S0140-6736(19)32316-5.

2. Song, J., Bergen, S.E., Kuja-Halkola, R., Larsson, H., Landén, M., and Lichtenstein, P. (2015). Bipolar disorder and its relation to major psychiatric disorders: a family-based study in the Swedish population. Bipolar Disord. 17, 184–193. 10.1111/bdi.12242.

3. Owen, M.J., Sawa, A., and Mortensen, P.B. (2016). Schizophrenia. The Lancet 388, 86–97. 10.1016/S0140-6736(15)01121-6.

4. Mullins, N., Forstner, A.J., O’Connell, K.S., Coombes, B., Coleman, J.R.I., Qiao, Z., Als, T.D., Bigdeli, T.B., Børte, S., Bryois, J., et al. (2021). Genome-wide association study of more than 40,000 bipolar disorder cases provides new insights into the underlying biology. Nat. Genet. 53, 817–829. 10.1038/s41588-021-00857-4.

5. Gibson, G. (2012). Rare and common variants: twenty arguments. Nat. Rev. Genet. 13, 135–145. 10.1038/nrg3118.

6. The Schizophrenia Psychiatric Genome-Wide Association Study (GWAS) Consortium (2011). Genome-wide association study identifies five new schizophrenia loci. Nat. Genet. 43, 969–976. 10.1038/ng.940.

7. Ripke, S., O’Dushlaine, C., Chambert, K., Moran, J.L., Kähler, A.K., Akterin, S., Bergen, S.E., Collins, A.L., Crowley, J.J., Fromer, M., et al. (2013). Genome-wide association analysis identifies 13 new risk loci for schizophrenia. Nat. Genet. 45, 1150–1159. 10.1038/ng.2742.

8. Schizophrenia Working Group of the Psychiatric Genomics Consortium (2014). Biological insights from 108 schizophrenia-associated genetic loci. Nature 511, 421–427. 10.1038/nature13595.

9. Pardiñas, A.F., Holmans, P., Pocklington, A.J., Escott-Price, V., Ripke, S., Carrera, N., Legge, S.E., Bishop, S., Cameron, D., Hamshere, M.L., et al. (2018). Common schizophrenia alleles are enriched in mutation-intolerant genes and in regions under strong background selection. Nat. Genet. 50, 381–389. 10.1038/s41588-018-0059-2.

10. Lam, M., Chen, C.-Y., Li, Z., Martin, A.R., Bryois, J., Ma, X., Gaspar, H., Ikeda, M., Benyamin, B., Brown, B.C., et al. (2019). Comparative genetic architectures of schizophrenia in East Asian and European populations. Nat. Genet. 51, 1670–1678. 10.1038/s41588-019-0512-x.

11. Sklar, P., Ripke, S., Scott, L.J., Andreassen, O.A., Cichon, S., Craddock, N., Edenberg, H.J., Nurnberger, J.I., Rietschel, M., Blackwood, D., et al. (2011). Large-scale genome-wide association analysis of bipolar disorder identifies a new susceptibility locus near ODZ4. Nat. Genet. 43, 977–983. 10.1038/ng.943.

12. Stahl, E.A., Breen, G., Forstner, A.J., McQuillin, A., Ripke, S., Trubetskoy, V., Mattheisen, M., Wang, Y., Coleman, J.R.I., Gaspar, H.A., et al. (2019). Genome-wide association study identifies 30 loci associated with bipolar disorder. Nat. Genet. 51, 793–803. 10.1038/s41588-019-0397-8.

13. Trubetskoy, V., Pardiñas, A.F., Qi, T., Panagiotaropoulou, G., Awasthi, S., Bigdeli, T.B., Bryois, J., Chen, C.-Y., Dennison, C.A., Hall, L.S., et al. (2022). Mapping genomic loci implicates genes and synaptic biology in schizophrenia. Nature 604, 502–508. 10.1038/s41586-022-04434-5.

14. Psychiatric GWAS Consortium Bipolar Disorder Working Group, Sklar, P., Ripke, S., Scott, L.J., Andreassen, O.A., Cichon, S., Craddock, N., Edenberg, H.J., Nurnberger, J.I., Rietschel, M., et al. (2011). Large-scale genome-wide association analysis of bipolar disorder identifies a new susceptibility locus near ODZ4. Nat. Genet. 43, 977–983. 10.1038/ng.943.

15. Zeng, J., Xue, A., Jiang, L., Lloyd-Jones, L.R., Wu, Y., Wang, H., Zheng, Z., Yengo, L., Kemper, K.E., Goddard, M.E., et al. (2021). Widespread signatures of natural selection across human complex traits and functional genomic categories. Nat. Commun. 12, 1164. 10.1038/s41467-021-21446-3.

16. Singh, T., Poterba, T., Curtis, D., Akil, H., Al Eissa, M., Barchas, J.D., Bass, N., Bigdeli, T.B., Breen, G., Bromet, E.J., et al. (2022). Rare coding variants in ten genes confer substantial risk for schizophrenia. Nature 604, 509–516. 10.1038/s41586-022-04556-w.

17. Palmer, D.S., Howrigan, D.P., Chapman, S.B., Adolfsson, R., Bass, N., Blackwood, D., Boks, M.P.M., Chen, C.-Y., Churchhouse, C., Corvin, A.P., et al. (2022). Exome sequencing in bipolar disorder identifies AKAP11 as a risk gene shared with schizophrenia. Nat. Genet. 54, 541–547. 10.1038/s41588-022-01034-x.

18. Homann, O.R., Misura, K., Lamas, E., Sandrock, R.W., Nelson, P., McDonough, S.I., and DeLisi, L.E. (2016). Whole-genome sequencing in multiplex families with psychoses reveals mutations in the SHANK2 and SMARCA1 genes segregating with illness. Mol. Psychiatry 21, 1690–1695. 10.1038/mp.2016.24.

19. Khan, F.F., Melton, P.E., McCarthy, N.S., Morar, B., Blangero, J., Moses, E.K., and Jablensky, A. (2018). Whole genome sequencing of 91 multiplex schizophrenia families reveals increased burden of rare, exonic copy number variation in schizophrenia probands and genetic heterogeneity. Schizophr. Res. 197, 337–345. 10.1016/j.schres.2018.02.034.

20. Sul, J.H., Service, S.K., Huang, A.Y., Ramensky, V., Hwang, S.-G., Teshiba, T.M., Park, Y., Ori, A.P.S., Zhang, Z., Mullins, N., et al. (2020). Contribution of common and rare variants to bipolar disorder susceptibility in extended pedigrees from population isolates. Transl. Psychiatry 10, 74. 10.1038/s41398-020-0758-1.

21. Alkelai, A., Greenbaum, L., Docherty, A.R., Shabalin, A.A., Povysil, G., Malakar, A., Hughes, D., Delaney, S.L., Peabody, E.P., McNamara, J., et al. (2022). The benefit of diagnostic whole genome sequencing in schizophrenia and other psychotic disorders. Mol. Psychiatry 27, 1435–1447. 10.1038/s41380-021-01383-9.

22. Georgi, B., Craig, D., Kember, R.L., Liu, W., Lindquist, I., Nasser, S., Brown, C., Egeland, J.A., Paul, S.M., and Bućan, M. (2014). Genomic View of Bipolar Disorder Revealed by Whole Genome Sequencing in a Genetic Isolate. PLoS Genet. 10, e1004229. 10.1371/journal.pgen.1004229.

23. Mojarad, B.A., Yin, Y., Manshaei, R., Backstrom, I., Costain, G., Heung, T., Merico, D., Marshall, C.R., Bassett, A.S., and Yuen, R.K.C. (2021). Genome sequencing broadens the range of contributing variants with clinical implications in schizophrenia. Transl. Psychiatry 11, 84. 10.1038/s41398-021-01211-2.

24. Biernacka, J., Jenkins, G., McDonnell, S., Batzler, A., Sicotte, H., Fogarty, Z., Welkie, B., Baheti, S., Coombes, B., McElroy, S., et al. (2019). A whole genome sequencing study identifies a rare variant in ANK3 that may contribute to bipolar disorder. Eur. Neuropsychopharmacol. 29, S901.

25. Halvorsen, M., Huh, R., Oskolkov, N., Wen, J., Netotea, S., Giusti-Rodriguez, P., Karlsson, R., Bryois, J., Nystedt, B., Ameur, A., et al. (2020). Increased burden of ultra-rare structural variants localizing to boundaries of topologically associated domains in schizophrenia. Nat. Commun. 11, 1842. 10.1038/s41467-020-15707-w.

26. Popejoy, A.B., and Fullerton, S.M. (2016). Genomics is failing on diversity. Nature 538, 161–164. 10.1038/538161a.

27. Sirugo, G., Williams, S.M., and Tishkoff, S.A. (2019). The Missing Diversity in Human Genetic Studies. Cell 177, 26–31. 10.1016/j.cell.2019.02.048.

28. Martin, A.R., Kanai, M., Kamatani, Y., Okada, Y., Neale, B.M., and Daly, M.J. (2019). Clinical use of current polygenic risk scores may exacerbate health disparities. Nat. Genet. 51, 584–591. 10.1038/s41588-019-0379-x.

29. Mahajan, A., Wessel, J., Willems, S.M., Zhao, W., Robertson, N.R., Chu, A.Y., Gan, W., Kitajima, H., Taliun, D., Rayner, N.W., et al. (2018). Refining the accuracy of validated target identification through coding variant fine-mapping in type 2 diabetes. Nat. Genet. 50, 559–571. 10.1038/s41588-018-0084-1.

30. Spracklen, C.N., Horikoshi, M., Kim, Y.J., Lin, K., Bragg, F., Moon, S., Suzuki, K., Tam, C.H.T., Tabara, Y., Kwak, S.-H., et al. (2020). Identification of type 2 diabetes loci in 433,540 East Asian individuals. Nature 582, 240–245. 10.1038/s41586-020-2263-3.

31. Taliun, D., Harris, D.N., Kessler, M.D., Carlson, J., Szpiech, Z.A., Torres, R., Taliun, S.A.G., Corvelo, A., Gogarten, S.M., Kang, H.M., et al. (2021). Sequencing of 53,831 diverse genomes from the NHLBI TOPMed Program. Nature 590, 290–299. 10.1038/s41586-021-03205-y.

32. Sanders, S.J., Neale, B.M., Huang, H., Werling, D.M., An, J.-Y., Dong, S., Abecasis, G., Arguello, P.A., Blangero, J., Boehnke, M., et al. (2017). Whole genome sequencing in psychiatric disorders: the WGSPD consortium. Nat. Neurosci. 20, 1661–1668. 10.1038/s41593-017-0017-9.

33. Pato, M.T., Sobell, J.L., Medeiros, H., Abbott, C., Sklar, B.M., Buckley, P.F., Bromet, E.J., Escamilla, M.A., Fanous, A.H., Lehrer, D.S., et al. (2013). The genomic psychiatry cohort: Partners in discovery. Am. J. Med. Genet. B Neuropsychiatr. Genet. 162, 306–312. 10.1002/ajmg.b.32160.

34. Bigdeli, T.B., Genovese, G., Georgakopoulos, P., Meyers, J.L., Peterson, R.E., Iyegbe, C.O., Medeiros, H., Valderrama, J., Achtyes, E.D., Kotov, R., et al. (2020). Contributions of common genetic variants to risk of schizophrenia among individuals of African and Latino ancestry. Mol. Psychiatry 25, 2455–2467. 10.1038/s41380-019-0517-y.

35. Swerdlow, N.R., Gur, R.E., and Braff, D.L. (2015). Consortium on the Genetics of Schizophrenia (COGS) assessment of endophenotypes for schizophrenia: An introduction to this Special Issue of schizophrenia research. Schizophr. Res. 163, 9–16. 10.1016/j.schres.2014.09.047.

36. Smith, E.N., Bloss, C.S., Badner, J.A., Barrett, T., Belmonte, P.L., Berrettini, W., Byerley, W., Coryell, W., Craig, D., Edenberg, H.J., et al. (2009). Genome-wide association study of bipolar disorder in European American and African American individuals. Mol. Psychiatry 14, 755–763. 10.1038/mp.2009.43.

37. Nierenberg, A.A., Friedman, E.S., Bowden, C.L., Sylvia, L.G., Thase, M.E., Ketter, T., Ostacher, M.J., Leon, A.C., Reilly-Harrington, N., Iosifescu, D.V., et al. (2013). Lithium Treatment Moderate-Dose Use Study (LiTMUS) for Bipolar Disorder: A Randomized Comparative Effectiveness Trial of Optimized Personalized Treatment With and Without Lithium. Am J Psychiatry.

38. Sklar, P., Smoller, J.W., Fan, J., Ferreira, M.A.R., Perlis, R.H., Chambert, K., Nimgaonkar, V.L., McQueen, M.B., Faraone, S.V., Kirby, A., et al. (2008). Whole-genome association study of bipolar disorder. Mol. Psychiatry 13, 558–569. 10.1038/sj.mp.4002151.

39. Jun, G., Wing, M.K., Abecasis, G.R., and Kang, H.M. (2015). An efficient and scalable analysis framework for variant extraction and refinement from population-scale DNA sequence data. Genome Res. 25, 918–925. 10.1101/gr.176552.114.

40. Center for Statistical Genetics (2020). statgen: topmed variant calling. GitHub. https://github.com/statgen/topmed_variant_calling.

41. Zhang, F., Flickinger, M., Taliun, S.A.G., InPSYght Psychiatric Genetics Consortium, Abecasis, G.R., Scott, L.J., McCaroll, S.A., Pato, C.N., Boehnke, M., and Kang, H.M. (2020). Ancestry-agnostic estimation of DNA sample contamination from sequence reads. Genome Res. 30, 185–194. 10.1101/gr.246934.118.

42. Price, A.L., Weale, M.E., Patterson, N., Myers, S.R., Need, A.C., Shianna, K.V., Ge, D., Rotter, J.I., Torres, E., Taylor, K.D., et al. (2008). Long-Range LD Can Confound Genome Scans in Admixed Populations. Am. J. Hum. Genet. 83, 132–135. 10.1016/j.ajhg.2008.06.005.

43. Purcell, S., Neale, B., Todd-Brown, K., Thomas, L., Ferreira, M.A.R., Bender, D., Maller, J., Sklar, P., De Bakker, P.I.W., Daly, M.J., et al. (2007). PLINK: A Tool Set for Whole-Genome Association and Population-Based Linkage Analyses. Am. J. Hum. Genet. 81, 559–575. 10.1086/519795.

44. Rosenberg, N.A., Pritchard, J.K., Weber, J.L., Cann, H.M., Kidd, K.K., Zhivotovsky, L.A., and Feldman, M.W. (2002). Genetic Structure of Human Populations. 298.

45. Manichaikul, A., Mychaleckyj, J.C., Rich, S.S., Daly, K., Sale, M., and Chen, W.-M. (2010). Robust relationship inference in genome-wide association studies. Bioinformatics 26, 2867–2873. 10.1093/bioinformatics/btq559.

46. Alexander, D.H., Novembre, J., and Lange, K. (2009). Fast model-based estimation of ancestry in unrelated individuals. Genome Res. 19, 1655–1664. 10.1101/gr.094052.109.

47. The 1000 Genomes Project Consortium, Corresponding authors, Auton, A., Abecasis, G.R., Steering committee, Altshuler, D.M., Durbin, R.M., Abecasis, G.R., Bentley, D.R., Chakravarti, A., et al. (2015). A global reference for human genetic variation. Nature 526, 68–74. 10.1038/nature15393.

48. Fan, S., Spence, J.P., Feng, Y., Hansen, M.E.B., Terhorst, J., Beltrame, M.H., Ranciaro, A., Hirbo, J., Beggs, W., Thomas, N., et al. (2023). Whole-genome sequencing reveals a complex African population demographic history and signatures of local adaptation. Cell 186, 923–939.e14. 10.1016/j.cell.2023.01.042.

49. Cann, H.M., Toma, C. de, Cazes, L., Legrand, M.-F., Morel, V., Piouffre, L., Bodmer, J., Bodmer, W.F., Bonne-Tamir, B., Cambon-Thomsen, A., et al. (2002). A Human Genome Diversity Cell Line Panel. Science 296, 261–262. 10.1126/science.296.5566.261b.

50. Maples, B.K., Gravel, S., Kenny, E.E., and Bustamante, C.D. (2013). RFMix: A Discriminative Modeling Approach for Rapid and Robust Local-Ancestry Inference. Am. J. Hum. Genet. 93, 278–288. 10.1016/j.ajhg.2013.06.020.

51. Li, J.Z., Absher, D.M., Tang, H., Southwick, A.M., Casto, A.M., Ramachandran, S., Cann, H.M., Barsh, G.S., Feldman, M., Cavalli-Sforza, L.L., et al. (2008). Worldwide Human Relationships Inferred from Genome-Wide Patterns of Variation. Science 319, 1100–1104. 10.1126/science.1153717.

52. Wang, C., Zhan, X., Bragg-Gresham, J., Kang, H.M., Stambolian, D., Chew, E.Y., Branham, K.E., Heckenlively, J., The FUSION Study, Fulton, R., et al. (2014). Ancestry estimation and control of population stratification for sequence-based association studies. Nat. Genet. 46, 409–415. 10.1038/ng.2924.

53. Purcell, S., Cherny, S.S., and Sham, P.C. (2003). Genetic Power Calculator: design of linkage andassociation genetic mapping studies of complex traits. Bioinformatics 19, 149–150. 10.1093/bioinformatics/19.1.149.

54. Zhou, W., Nielsen, J.B., Fritsche, L.G., Dey, R., Gabrielsen, M.E., Wolford, B.N., LeFaive, J., VandeHaar, P., Gagliano, S.A., Gifford, A., et al. (2018). Efficiently controlling for case-control imbalance and sample relatedness in large-scale genetic association studies. Nat. Genet. 50, 1335–1341. 10.1038/s41588-018-0184-y.

55. Cross-Disorder Group of the Psychiatric Genomics Consortium (2013). Identification of risk loci with shared effects on five major psychiatric disorders: a genome-wide analysis. The Lancet 381, 1371–1379. 10.1016/S0140-6736(12)62129-1.

56. Zhan, X., Hu, Y., Li, B., Abecasis, G.R., and Liu, D.J. (2016). RVTESTS: an efficient and comprehensive tool for rare variant association analysis using sequence data. Bioinformatics 32, 1423–1426. 10.1093/bioinformatics/btw079.

57. Purcell, S.M., Moran, J.L., Fromer, M., Ruderfer, D., Solovieff, N., Roussos, P., O’Dushlaine, C., Chambert, K., Bergen, S.E., Kähler, A., et al. (2014). A polygenic burden of rare disruptive mutations in schizophrenia. Nature 506, 185–190. 10.1038/nature12975.

58. Genovese, G., Fromer, M., Stahl, E.A., Ruderfer, D.M., Chambert, K., Landén, M., Moran, J.L., Purcell, S.M., Sklar, P., Sullivan, P.F., et al. (2016). Increased burden of ultra-rare protein-altering variants among 4,877 individuals with schizophrenia. Nat. Neurosci. 19, 1433–1441. 10.1038/nn.4402.

59. Fromer, M., Pocklington, A.J., Kavanagh, D.H., Williams, H.J., Dwyer, S., Gormley, P., Georgieva, L., Rees, E., Palta, P., Ruderfer, D.M., et al. (2014). De novo mutations in schizophrenia implicate synaptic networks. Nature 506, 179–184. 10.1038/nature12929.

60. Singh, T., Walters, J.T.R., Johnstone, M., Curtis, D., Suvisaari, J., Torniainen, M., Rees, E., Iyegbe, C., Blackwood, D., McIntosh, A.M., et al. (2017). The contribution of rare variants to risk of schizophrenia in individuals with and without intellectual disability. Nat. Genet. 49, 1167–1173. 10.1038/ng.3903.

61. Vu, H., and Ernst, J. (2022). Universal annotation of the human genome through integration of over a thousand epigenomic datasets. Genome Biol. 23, 9. 10.1186/s13059-021-02572-z.

62. Arneson, A., and Ernst, J. (2019). Systematic discovery of conservation states for single-nucleotide annotation of the human genome. Commun. Biol. 2, 248. 10.1038/s42003-019-0488-1.

63. Arneson, A., Felsheim, B., Chien, J., and Ernst, J. (2020). ConsHMM Atlas: conservation state annotations for major genomes and human genetic variation. NAR Genomics Bioinforma. 2, lqaa104. 10.1093/nargab/lqaa104.

64. Amemiya, H.M., Kundaje, A., and Boyle, A.P. (2019). The ENCODE Blacklist: Identification of Problematic Regions of the Genome. Sci. Rep. 9, 9354. 10.1038/s41598-019-45839-z.

65. He, Z., Xu, B., Buxbaum, J., and Ionita-Laza, I. (2019). A genome-wide scan statistic framework for whole-genome sequence data analysis. Nat. Commun. 10, 3018. 10.1038/s41467-019-11023-0.

66. Karolchik, D. (2003). The UCSC Genome Browser Database. Nucleic Acids Res. 31, 51–54. 10.1093/nar/gkg129.

67. Benson, G. (1999). Tandem repeats finder: a program to analyze DNA sequences. Nucleic Acids Res. 27, 573–580. 10.1093/nar/27.2.573.

68. Ernst, J., and Kellis, M. (2015). Large-scale imputation of epigenomic datasets for systematic annotation of diverse human tissues. Nat. Biotechnol. 33, 364–376. 10.1038/nbt.3157.

69. Lu, J.Y., Shao, W., Chang, L., Yin, Y., Li, T., Zhang, H., Hong, Y., Percharde, M., Guo, L., Wu, Z., et al. (2020). Genomic Repeats Categorize Genes with Distinct Functions for Orchestrated Regulation. Cell Rep. 30, 3296–3311.e5. 10.1016/j.celrep.2020.02.048.

70. Tom, J.A., Reeder, J., Forrest, W.F., Graham, R.R., Hunkapiller, J., Behrens, T.W., and Bhangale, T.R. (2017). Identifying and mitigating batch effects in whole genome sequencing data. BMC Bioinformatics 18, 351. 10.1186/s12859-017-1756-z.

71. Werling, D.M., Brand, H., An, J.-Y., Stone, M.R., Zhu, L., Glessner, J.T., Collins, R.L., Dong, S., Layer, R.M., Markenscoff-Papadimitriou, E., et al. (2018). An analytical framework for whole-genome sequence association studies and its implications for autism spectrum disorder. Nat. Genet. 50, 727–736. 10.1038/s41588-018-0107-y.

72. Robinson, N., and Bergen, S.E. (2021). Environmental Risk Factors for Schizophrenia and Bipolar Disorder and Their Relationship to Genetic Risk: Current Knowledge and Future Directions. Front. Genet. 12, 686666. 10.3389/fgene.2021.686666.

73. Legge, S.E., Pardiñas, A.F., Woolway, G., Rees, E., Cardno, A.G., Escott-Price, V., Holmans, P., Kirov, G., Owen, M.J., O’Donovan, M.C., et al. (2024). Genetic and Phenotypic Features of Schizophrenia in the UK Biobank. JAMA Psychiatry 81, 681. 10.1001/jamapsychiatry.2024.0200.

74. Moskvina, V., Holmans, P., Schmidt, K.M., and Craddock, N. (2005). Design of Case-controls Studies with Unscreened Controls. Ann. Hum. Genet. 69, 566–576. 10.1111/j.1529-8817.2005.00175.x.

