## Supplemental information for "Whole genome sequence-based association analysis of African American individuals with bipolar disorder and schizophrenia"

#### Supplemental Text

*Follow-up analyses for the top window across all case-control test combinations*

We conducted follow-up analyses for the top window across all six case-control test combinations, that is chr11:64,859,972-64,869,939 ($p=4.06 \times 10^{-8}$ with repeats and $p=1.12 \times 10^{-9}$ without repeats for InPSYght BD vs. InPSYght controls) (**Figure S13A, B**). We first analyzed the extent to which the most significant variants could explain the overall association signal. To do this we first performed single-variant two-sided Fisher's exact test (Methods) on each of the 197 variants not overlapping repeats in the region for InPSYght BD cases versus controls. The 12 most significant variants in this region had p-values within range [$1.5 \times 10^{-4}$, $1.5 \times 10^{-2}$] while no other individual variant was marginally significant (p-value <0.05). 11 out of these 12 variants are noncoding, with chr11:64859972G,A being a synonymous coding variant. When we restricted the burden test to only these 12 variants, we observed a p-value of $9.0 \times 10^{-8}$ whereas performing the same test on all other variants in the window only yields a p-value of $4.8 \times 10^{-3}$, indicating a major contribution from the 12 nominally significant variants. These variants constitute two distinct haplotypes, both of which being abundant among samples African American individuals (**Figure S13C**).

The *EHD1* gene which overlaps the significant window was previously shown to play potential roles in neuropsychiatric disorders. Earlier studies established EHD1 as mediators of synaptic exo- and endocytosis[1–3]. A small-sample WES study for BD trios identified a *de novo* frame-shift mutation within the *EHD1* gene[3] and a follow-up knock-in mice study suggested that this mutation induces BD-like behavior in mouse models[4].

In addition, the significant window was notable in that it sits in a region of the genome that is dense for common variants based on the most recent PGC BD GWAS study of common variants[5]. Three distinct common variants associated within BD are within 2 Mb and five are within 6 Mb (**Table S9**). Relative to other 10kb windows in the genome this window was in the top 0.037% and 0.097% in terms of the maximum distance to the third and fifth closest variants respectively. The nearest lead variant at genome-wide significant loci for BD was 622kb away. Interestingly the window overlaps a near-significant loci (p-value ~$7.0 \times 10^{-8}$) in the most recent PGC SZ GWAS study[6], while the closest genome-wide significant loci for SZ (p-value =$1.8 \times 10^{-9}$) is 0.8 Mb away (**Figure S14B**).

We also investigated the chromatin state context of the window. We used chromatin state annotations of the genome for 127 cell and tissue types based on a 25-state ChromHMM model previously learned from 12 marks based on imputed data[7,8]. This region was notable in that the prevalence of the TxReg (transcribed and regulatory) state, which is associated with concurrently transcribed, promoter and enhancer chromatin marks, broadly across the 10kb window and across cell and tissue types (**Figure S14A, S14B**). On average 44.5% of bases in the 10kb window were assigned to the TxReg state in a cell or tissue type. The next most prevalent state in the window was TxEnh5' which covered 29.6% of bases on average and shared high levels of enhancer marks and some transcription marks with the TxReg state. To quantify how extreme the prevalence of these states are in the window relative to the rest of the genome, for each of the 10kb windows throughout the genome we computed the percentage of 200 base pair bins across all cell and tissue types that are assigned to each state. The significant window ranked in the top 0.02% of all 10kb windows in the genome based on state TxReg and the top 0.08% based on TxEnh5' (**Figure S14C**). These results motivated us to inspect the most extreme locations in the genome in terms of the density of TxReg. Notably, the top 1st and 3rd ranking windows that represent unique loci are only less than 0.6 Mb away and overlap with the transcriptional sites of the long noncoding RNAs (lncRNAs) NEAT1 (chr11: 65,422,774-65,445,540) and MALAT1 (chr11: 65,497,688-65,506,516) respectively, both of which are distinguished among lncRNAs in their broad binding across the genome[9]. Notably, as indicated by GRID-seq (capturing of global RNA interactions with DNA by deep sequencing) data on the MM.1S cell line[9], the significant window resides in a broader region enriched for both *NEAT1* and *MALAT1* binding sites (**Figure S14B**). There has been emerging evidence linking lncRNAs to various brain disorders[10,11]. Specifically, studies have shown that NEAT1 is down-regulated in SZ[12] while *MALAT1* is down-regulated in BD[13].

#### Supplemental Tables

**Table S1**. InPSYght study samples by sub-cohort and case/control status

| Study of origin | BD (N) | SZ (N) | Control (N) |
| --- | --- | --- | --- |
| GPC | 1475 | 2855 | 2338 |
| COGS | 0 | 432 | 313 |
| NIMH Repository: (STEP-BD, BIGS, Litmus) | 123 | 8 | 0 |

**Table S2**. Source of TOPMed controls from 11 TOPMed studies

| TOPMed study | Samples used as controls (N) | Sequencing Center |
|---|---|---|
| Cleveland Family Study (sleep apnea) | 2 | University of Washington (UW) |
| Barbados Asthma Genetics Study (BAGS) | 997 | Illumina |
| Genetic Epidemiology of COPD | 3171 | Broad |
| Multi-Ethnic Study of Atherosclerosis (MESA) | 1207 | Broad |
| African American Coronary Artery Calcification (AACAC); substudy of MESA | 127 | Broad |
| Atherosclerosis Risk in Communities (ARIC+VTE) | 295 | Baylor |
| Women's Health Initiative (WHI) | 1455 | Broad |
| Hypertension Genetic Epidemiology Network (HyperGen/GENOA) | 2988 | University of Washington (UW) |
| Sarcoidosis in African Americans | 632 | Baylor |
| BioMe Biobank | 1360 | Washington University (WashU) |
| CARDIA | 1578 | Baylor |

**Table S3.** InPSYght sample demographic and sequencing wave characteristics

| | SZ | BD | SZ + BD | Control | TOTAL |
|---|---|---|---|---|---|
| **N** | 3295 | 1598 | 4893 | 2651 | 7544 |
| **Females N (%)** | 1036 (31%) | 771 (48%) | 1807 (34%) | 1352 (51%) | 3159 (42%) |
| **Age Years (SD) (N non-missing)** | 43.7 (12.2) (3236) | 43.1 (11.7) (1521) | 43.5 (12.0) (4757) | 40.6 (13.6) (2631) | 42.5 (12.7) (7388) |
| **Sequencing Wave 0** | 112 | 5 | 117 | 114 | 231 |
| **1** | 165 | 165 | 330 | 161 | 491 |
| **2** | 161 | 165 | 326 | 165 | 491 |
| **3** | 599 | 118 | 717 | 344 | 1061 |
| **4** | 1019 | 223 | 1242 | 516 | 1758 |
| **5** | 795 | 512 | 1307 | 744 | 2051 |
| **6** | 444 | 410 | 854 | 607 | 1461 |

**Table S4.** InPSYght sample quality control exclusions

| Quality control | Individuals removed (N) |
|---|---|
| Duplicates | 366 |
| Sex mismatches | 20 |
| Non-XX/XY sex karyotypes | 17 |
| DNA contamination >5% | 4 |
| <98% of sites not at sequencing depth of 10 | 14 |
| Unqualified controls | 10 |

**Table S5**. Descriptions of the sequencing quality metrics and their loadings in the first four sequencing metadata PCs

| Metric | PC1 Loading | PC2 Loading | PC3 Loading | PC4 Loading | Description |
|---|---|---|---|---|---|
| VB_DEPTH | 0.70 | 0.72 | 0.02 | 0.03 | Average depth estimated by verifyBamID2 (in array genotyped sites) |
| FREEMIX | 0.00 | 0.00 | 0.00 | -0.01 | Contamination estimates by verifyBamID2 |
| CONTAM_PC1 | 0.00 | 0.00 | 0.00 | 0.01 | Estimated ancestry PC1,2,3,4 of contaminating samples estimated by verifyBamID2 |
| CONTAM_PC2 | 0.00 | 0.00 | 0.01 | -0.04 | |
| CONTAM_PC3 | 0.00 | 0.00 | 0.00 | 0.00 | |
| CONTAM_PC4 | 0.00 | 0.00 | 0.00 | 0.00 | |
| FRAC_DP1 | 0.00 | 0.00 | 0.00 | 0.00 | Fraction of bases covered by depths 1,5,10,15,20,30 |
| FRAC_DP5 | 0.00 | 0.00 | 0.00 | 0.00 | |
| FRAC_DP10 | 0.00 | 0.00 | 0.00 | -0.01 | |
| FRAC_DP15 | 0.00 | 0.00 | 0.00 | -0.11 | |
| FRAC_DP20 | 0.01 | 0.00 | 0.01 | -0.52 | |
| FRAC_DP30 | 0.03 | 0.01 | 0.03 | -0.84 | |
| RDP_X | 0.00 | -0.02 | 0.77 | 0.02 | Relative depth of X (compared to autosomes) |
| RDP_Y | 0.00 | 0.02 | -0.64 | -0.02 | Relative depth of Y (compared to autosomes) |
| AUTO_DP | 0.72 | -0.70 | -0.02 | 0.01 | Autosome depth |

**Table S6.** Summary of single-variant GWAS results. Significant threshold set to p<5E-9. Variant called on GRCh38.

| Sample group 1 | Sample group 2 | Genomic inflation factor (λ) | Number of significant loci (variant(s)). | Notes |
|---|---|---|---|---|
| InPSYght controls | TOPMed controls | 1.02 | 1 (chr13:79615934:CT:C) | Variant failed SVM filtering pipeline in TOPMed previous freeze; just passed filters in freeze9 (freeze used in current paper) |
| InPSYght BD + SZ | InPSYght + TOPMed controls | 1.01 | 0 (0) | |
| | InPSYght controls | 0.996 | 0 (0) | |
| InPSYght BD | InPSYght + TOPMed controls | 1.02 | 1 (chr18:49738979:G:T; chr18:49741194:A:G; chr18:49744269:GGT:G) | No longer significant with inclusion of meta-PCs |
| | InPSYght controls | 0.996 | 0 (0) | |
| InPSYght SZ | InPSYght + TOPMed controls | 1.00 | 0 (0) | |
| | InPSYght controls | 0.997 | 0 (0) | |

**Table S7**. Non-reference allele frequencies of the 3 significant chromosome 18 variants (InPSYght bipolar versus InPSYght + TOMed controls single-variant association analysis) in cases, controls and the total TOPMed study *Obtained from files from the download page of https://imputationserver.sph.umich.edu/rsq-browser/

| chr:bp:ref:alt (GRCh38) | rsID | Alternate allele frequency | | | | | | |
|---|---|---|---|---|---|---|---|---|
| | | InPSYght | | | TOPMed | InPSYght +TOPMed | Total TOPMed | |
| | | BD | SZ | Controls | Controls | Controls | AFR* | EUR* |
| Chr18: 49738979:G:T | rs796645723 | 0.0069 | 0.0014 | 0.00057 | 0.0012 | 0.0011 | 0.0033 | n/a |
| Chr18: 49741194:A:G | rs566405039 | 0.0069 | 0.0014 | 0.00075 | 0.0012 | 0.0011 | 0.0033 | n/a |
| Chr18: 49744269:GGT:G | rs796880023 | 0.0069 | 0.0014 | 0.00057 | 0.0012 | 0.0011 | 0.0031 | n/a |

**Table S8.** Comparison of singleton PTV burden test results from top ten implicated schizophrenia genes identified in SCHEMA.

| Gene | InPSYght (P-value) | SCHEMA OR (95% CI) | SCHEMA (P-value) |
|---|---|---|---|
| *SETD1A* | 0.86 | 20.1 (5.68-108) | 2.00e-12 |
| *CUL1* | *NA* | 36.1 (5.01-1570) | 2.01e-09 |
| *XPO7* | 0.86 | 52.2 (7.84-2190) | 7.18e-09 |
| *TRIO* | 0.89 | 5.02 (2.47-10.4) | 6.35e-08 |
| *CACNA1G* | 0.85 | 3.09 (1.21-7.63) | 4.57e-07 |
| *SP4* | *NA* | 9.37 (3.38-29.7) | 5.08e-07 |
| *GRIA3* | *NA* | Inf (4.73-Inf) | 5.98e-07 |
| *GRIN2A* | *NA* | 18.1 (3.74-172) | 7.37e-07 |
| *HERC1* | 0.31 | 3.51 (2.04-6.03) | 1.26e-06 |
| *RB1CC1* | 0.53 | 10 (2.89-43.9) | 2.00e-06 |

**Table S9**. Distances from the significant chromosome 11 window (chr11:64,859,972-64,869,939) to the closest GWAS loci reported in a PGC BD GWAS study[5], and the percentage of all 10kb genomic windows with smaller distance to their closest GWAS loci.

| Nth closest GWAS locus | Distance to GWAS locus (bp) | Percentage of 10kb genomic windows with smaller distance to its Nth closest GWAS loci |
|---|---|---|
| 1 | 622,550 | 2.59% |
| 2 | 1,216,310 | 0.47% |
| 3 | 1,692,155 | 0.037% |
| 4 | 3,013,821 | 0.046% |
| 5 | 5,806,865 | 0.097% |
| 6 | 14,516,526 | 0.40% |

#### Supplemental Figures

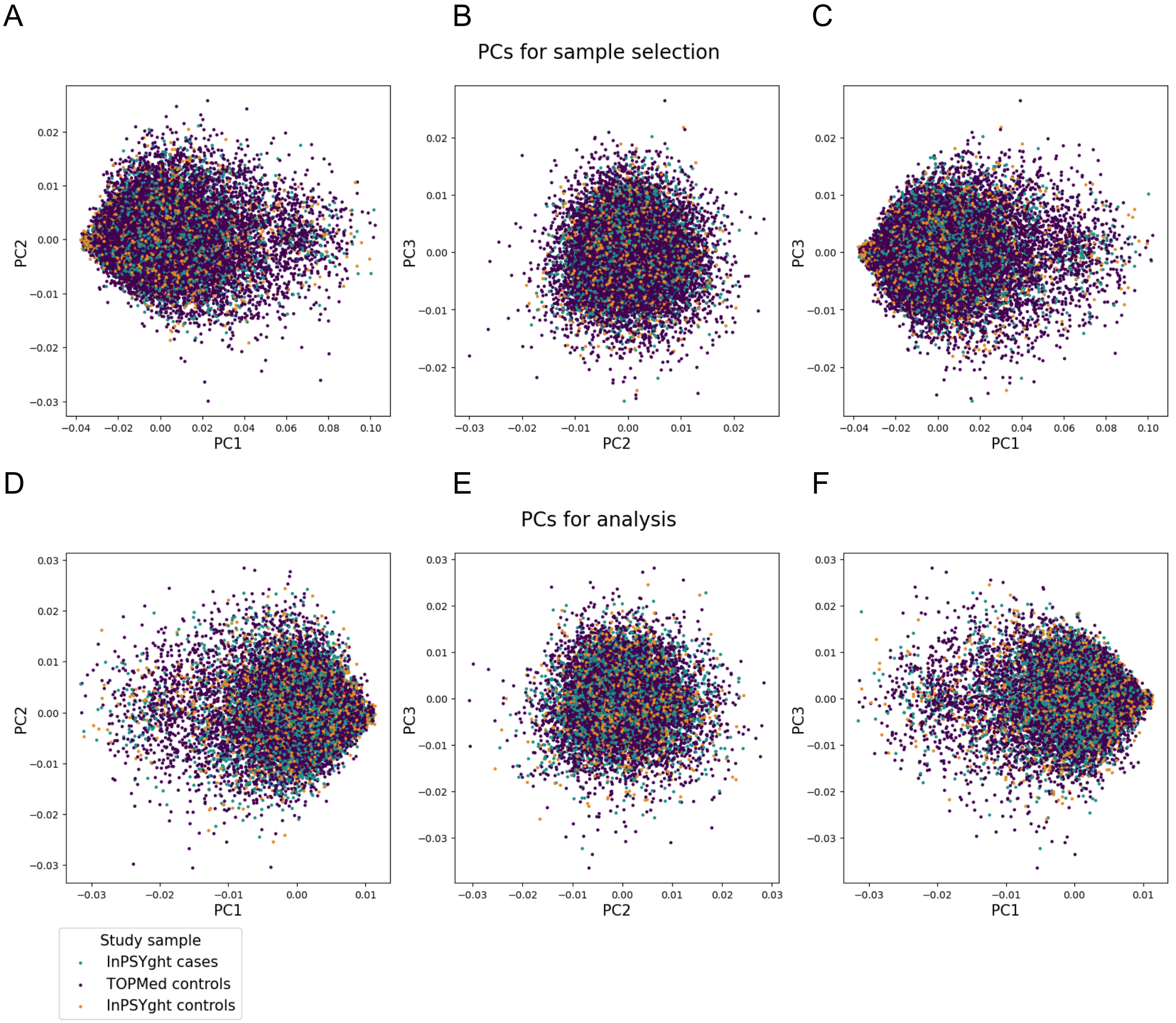


**Figure S1**. The first three genetic principal components (PCs) for the InPSYght and TOPMed samples. (A-C) PCs used for sample selection. (A) PC1 vs. PC2 (B) PC2 vs. PC3 and (C) PC1 vs. PC3. (D-F) PCs used for analysis. (D) PC1 vs. PC2 (E) PC2 vs. PC3 and (F) PC1 vs. PC3.

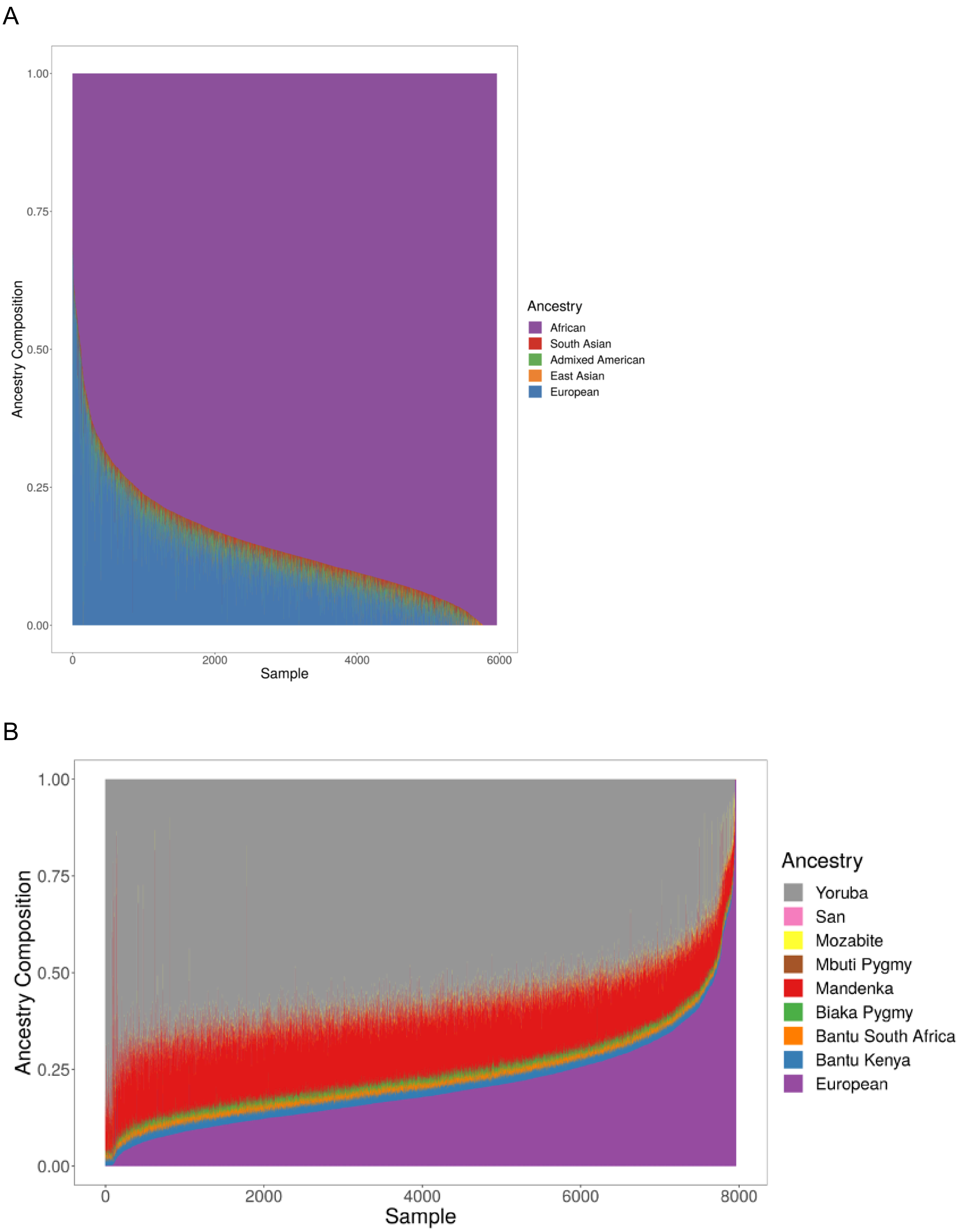
A
Ancestry Composition
1.00
0.75
0.50
0.25
0.00
0
2000
4000
6000
Sample
Ancestry
African
South Asian
Admixed American
East Asian
European
B
Ancestry Composition
1.00
0.75
0.50
0.25
0.00
0
2000
4000
6000
8000
Sample
Ancestry
Yoruba
San
Mozabite
Mbuti Pygmy
Mandenka
Biaka Pygmy
Bantu South Africa
Bantu Kenya
European

**Figure S2**. Estimated African and European ancestry composition of African American individuals within the InPSYght study. Global ancestry composition of African American individuals within the InPSYght study using 1000 Genomes super-populations (A) and within African sub-populations (B) Reference populations include European as a super population and African sub populations from the HGDP reference panel. Note this ancestry analysis differs from the 1000 Genomes Project phase 3 ancestry super-populations ancestry estimation analysis from which proportion European ancestry was estimated for InPSYght study inclusion. The 1000 Genomes Project based analysis including Admixed Americans (AMR), South Asian (SAS), and East Asian (EAS) individuals.

**A**

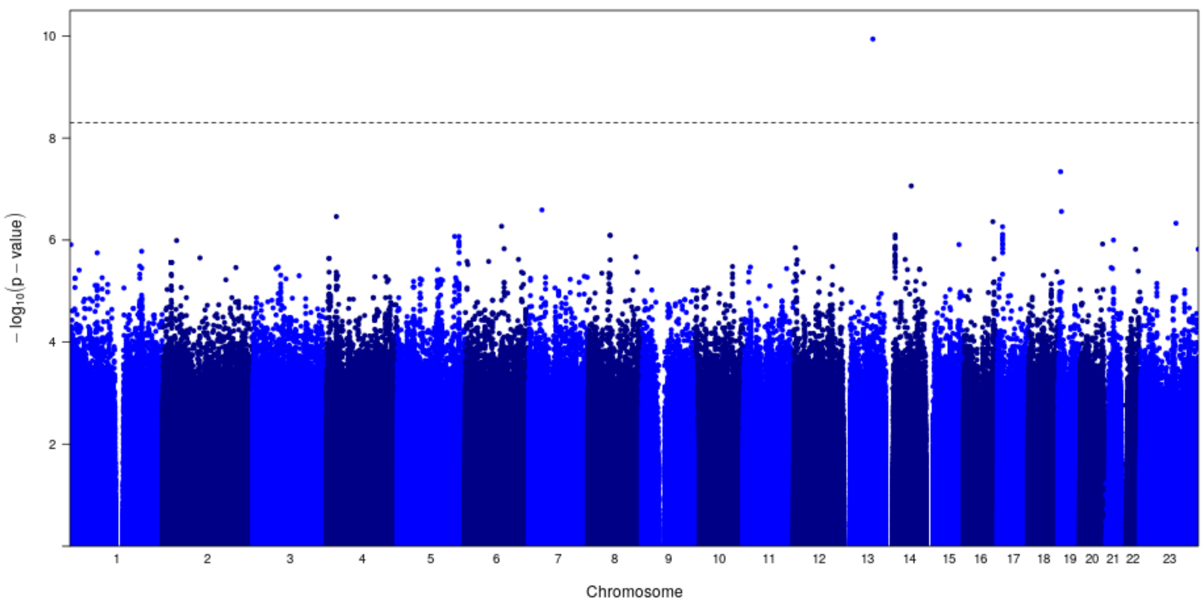


**B**

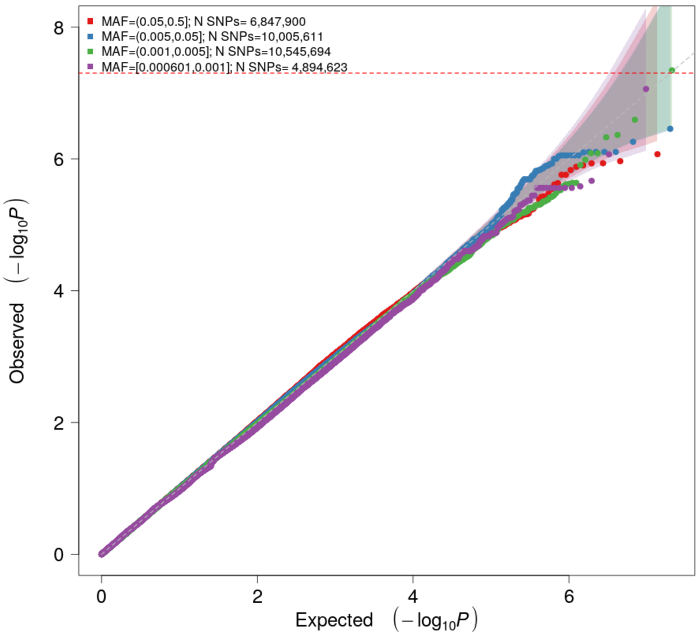


**Figure S3.** InPSYght controls versus TOPMed controls with sex and 10 PCs as covariates stratified by MAF of variants: Manhattan (A) and Quantile-Quantile Plots (B). Lambda GC: 1.02. Horizontal line shows genome-wide significance threshold of 5 x $10^{-9}$.

A

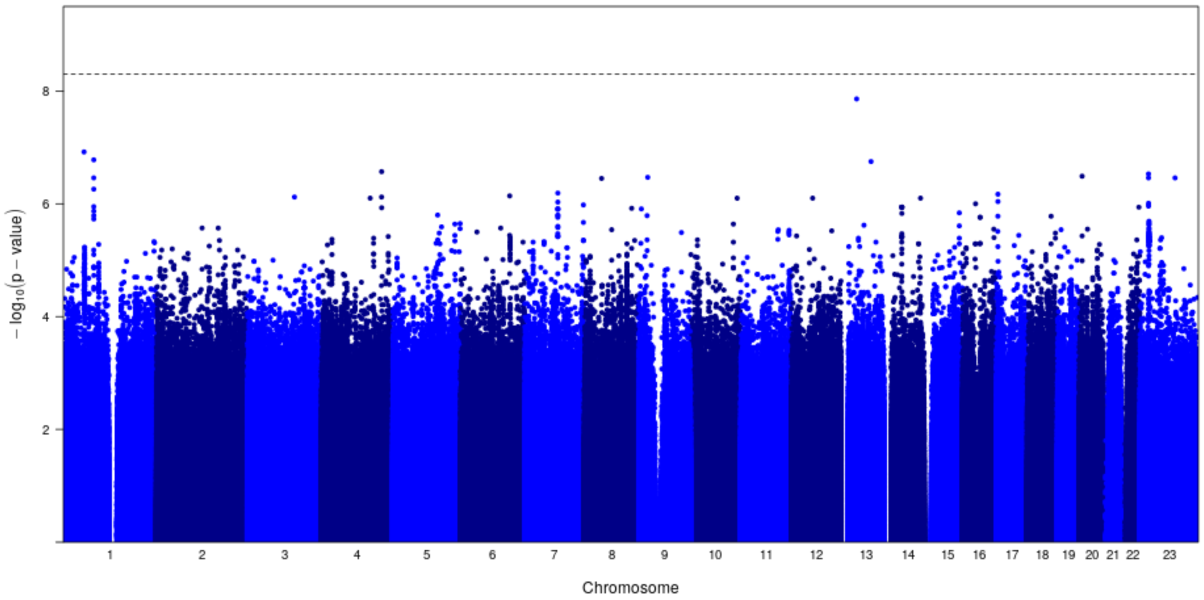


B

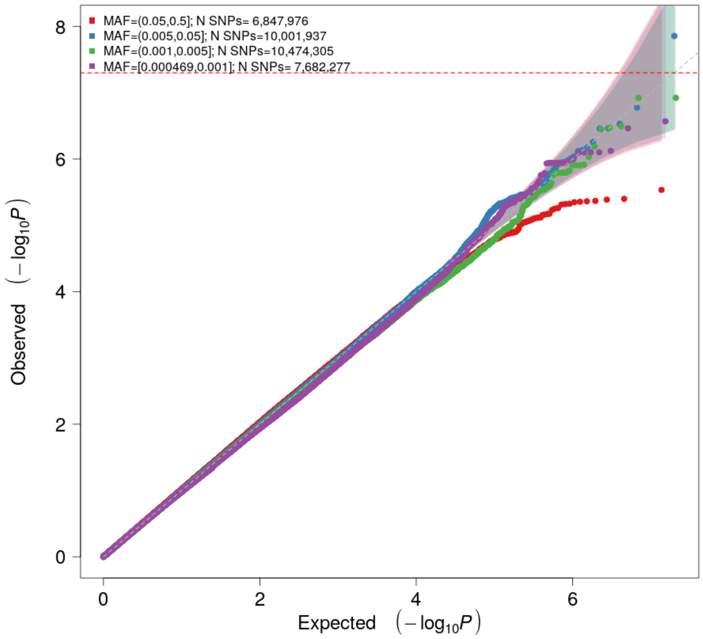


**Figure S4.** InPSYght cases with InPSYght + TOPMed controls with sex and 10 PCs as covariates stratified by MAF of variants: Manhattan (A) and Quantile-Quantile Plots (B). Lambda GC: 1.01. Horizontal line shows genome-wide significance threshold of 5 x $10^{-9}$.

A

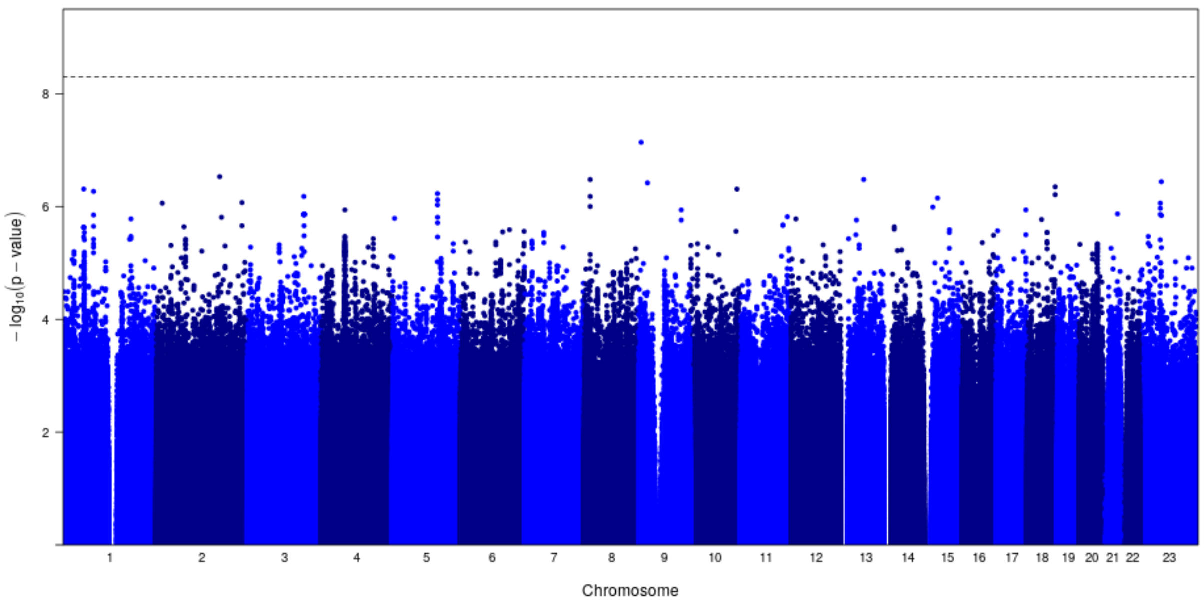


B

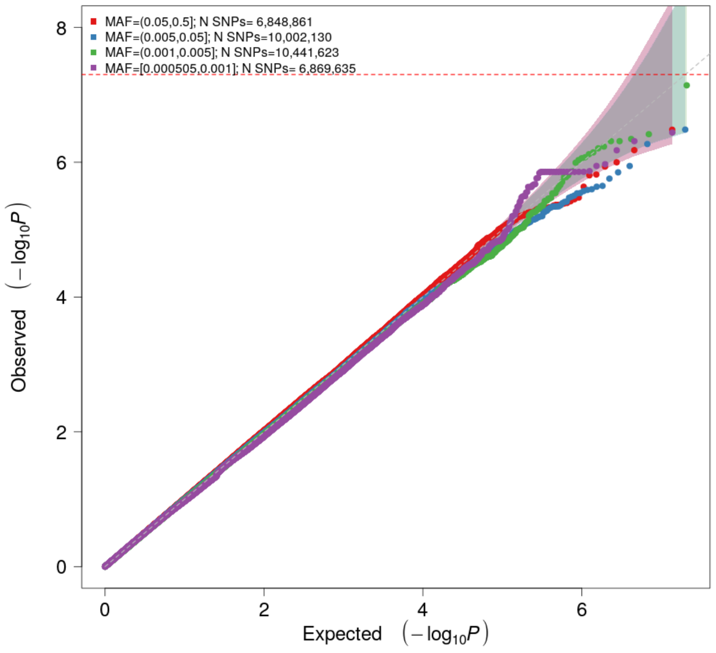


**Figure S5.** InPSYght schizophrenia cases versus InPSYght + TOPMed controls with sex and 10 PCs as covariates stratified by MAF of variants: Manhattan (A) and Quantile-Quantile Plots (B). Lambda GC: 1.00. Horizontal line shows genome-wide significance threshold of 5 x $10^{-9}$.

A

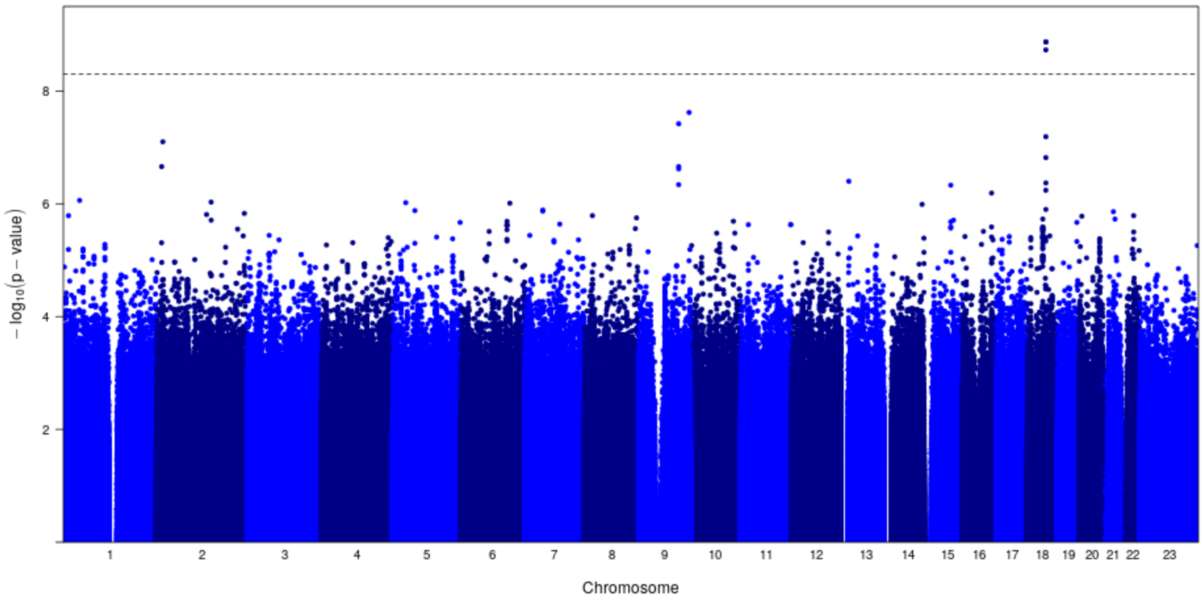


B

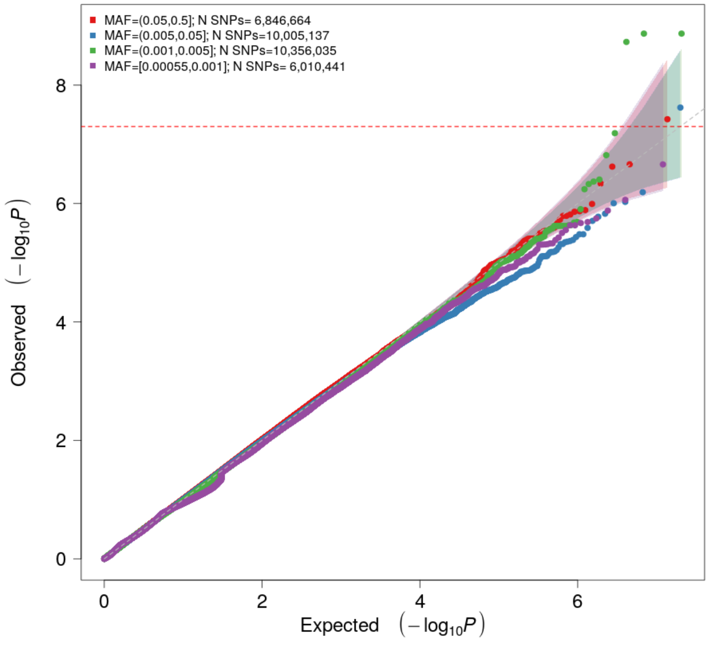


**Figure S6.** InPSYght bipolar cases versus InPSYght + TOPMed controls with sex and 10 PCs as covariates stratified by MAF of variants: Manhattan (A) and Quantile-Quantile Plots (B). Lambda GC: 1.02. Note: the Manhattan (Figure S6a is also presented as a main figure.) Horizontal line shows genome-wide significance threshold of 5 x $10^{-9}$.

**A**

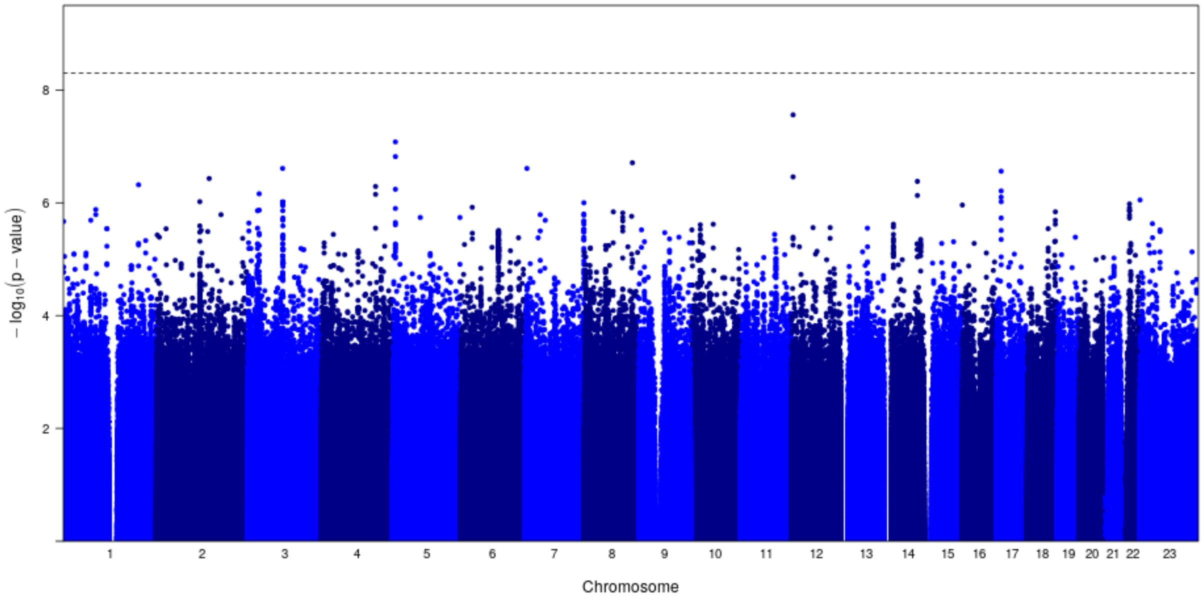


**B**

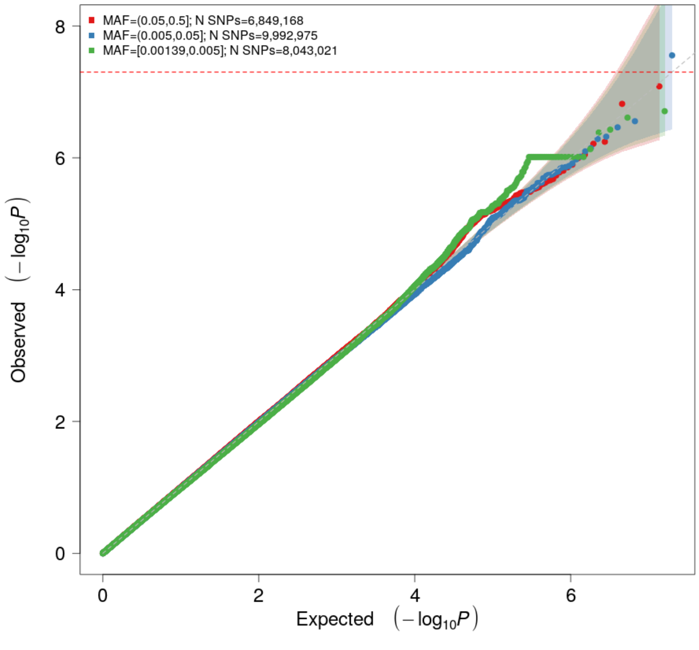


**Figure S7.** InPSYght-only case-control with sex, batch and 10 PCs as covariates stratified by MAF of variants: Manhattan (A) and Quantile-Quantile Plots (B). Lamb GC: 1.00. Horizontal line shows genome-wide significance threshold of 5 x $10^{-9}$.

**A**

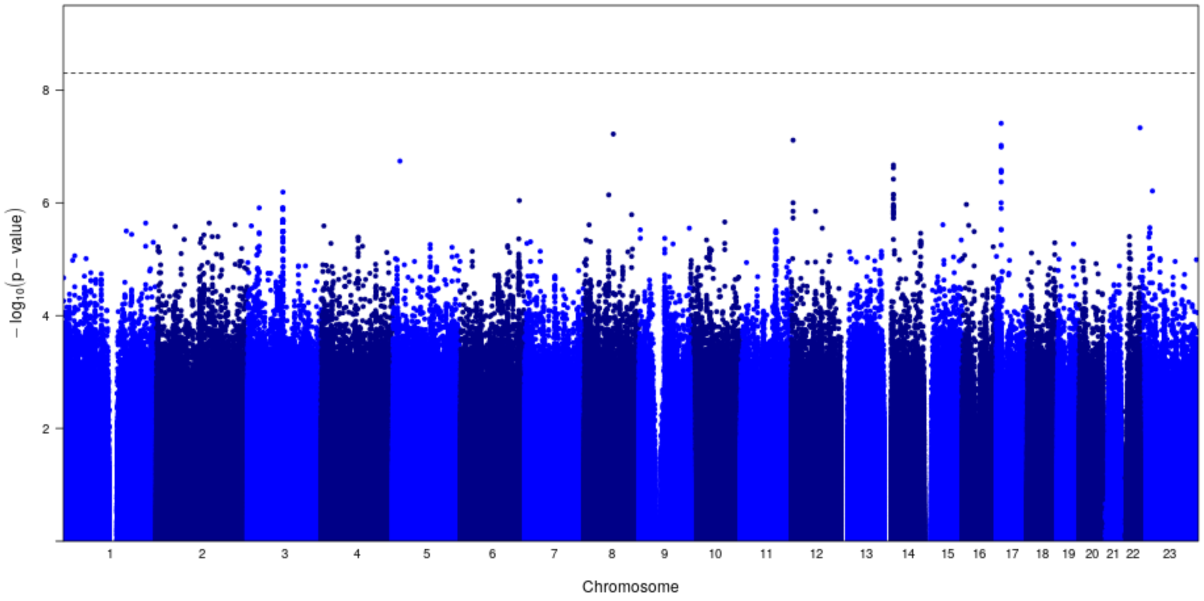


**B**

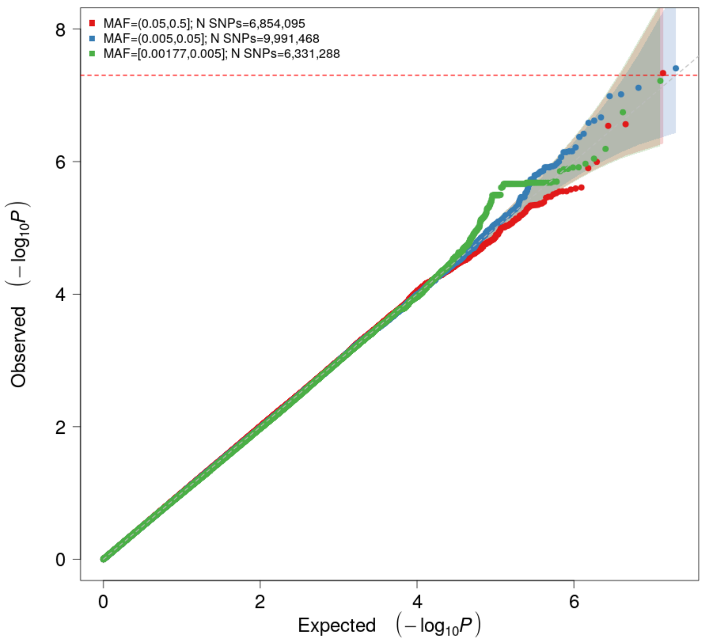


**Figure S8.** InPSYght-only SZ-control with sex, batch and 10 PCs as covariates stratified by MAF of variants: Manhattan (A) and Quantile-Quantile Plots (B). Lambda GC: 1.00. Horizontal line shows genome-wide significance threshold of 5 x $10^{-9}$.

**A**

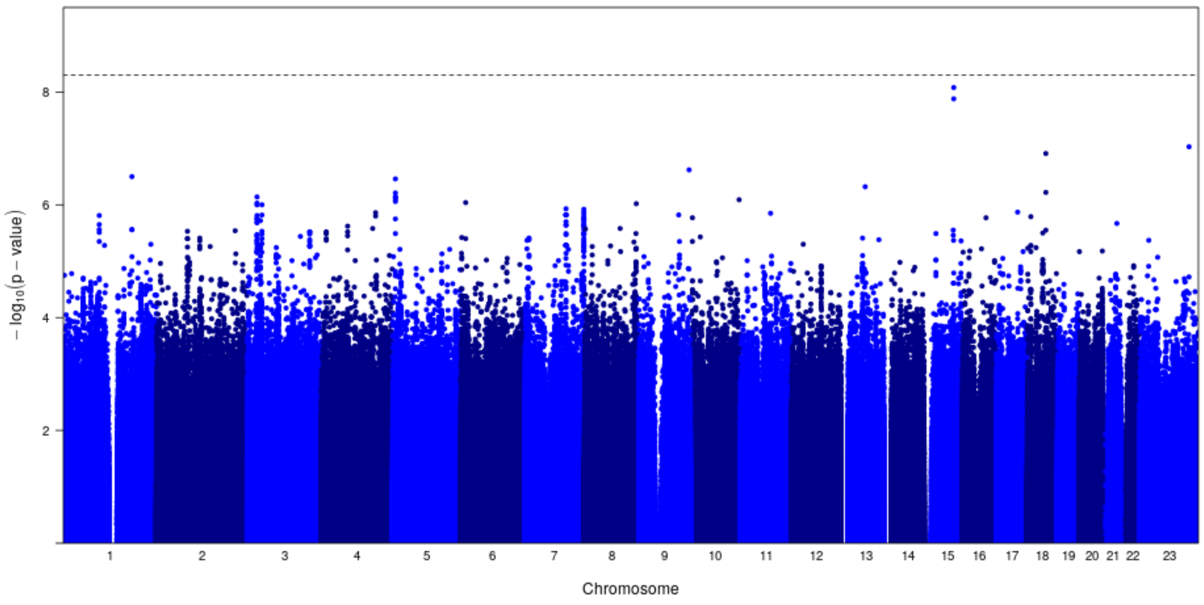


**B**

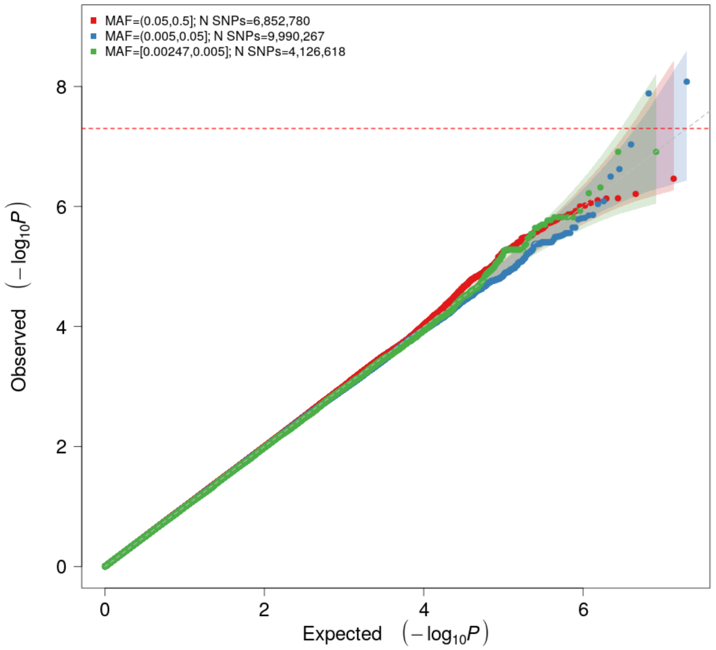


**Figure S9**. InPSYght-only bipolar-control with sex, batch and 10 PCs as covariates stratified by MAF of variants: Manhattan (A) and Quantile-Quantile Plots (B). Lambda GC: 1.00. Horizontal line shows genome-wide significance threshold of 5 x $10^{-9}$.

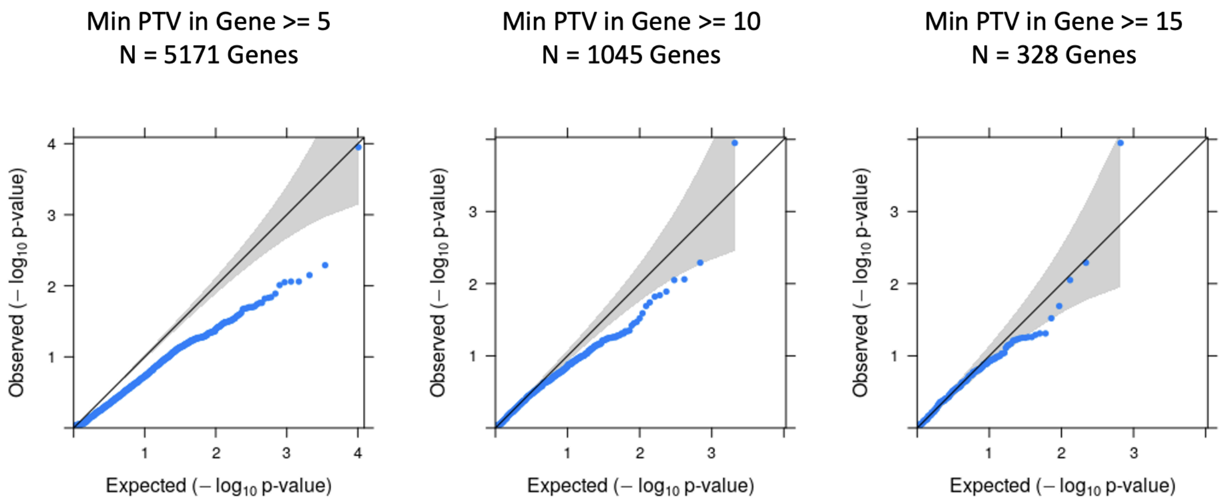


**Figure S10**. QQ plot of genome wide gene-level singleton PTV burden tests in unrelated individuals by minimum number of PTV criteria for gene inclusion. Covariates adjusted for include genetic PC1-10, sex, and total number of singletons.

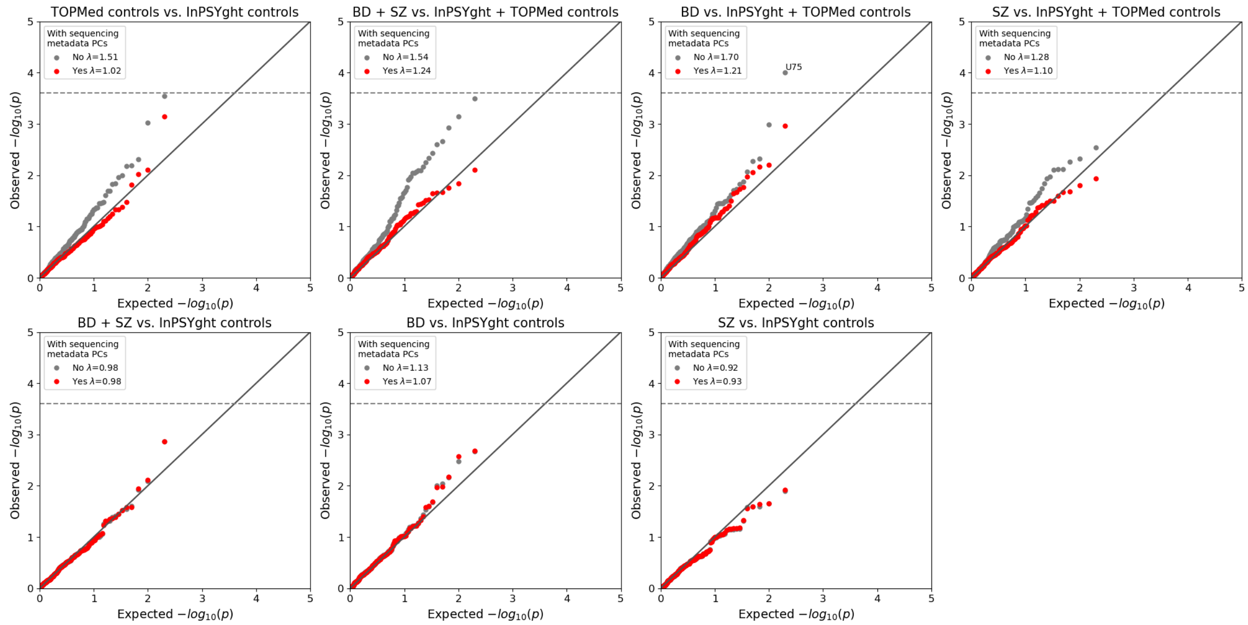


**Figure S11**. QQ-plots of chromatin and conservation state burden test results for different combinations of samples, showing their genomic inflation factors (λ) before and after including sequencing metadata PCs in the covariates. Horizontal lines show Bonferroni-based p-value thresholds (p=0.05/200). Diagonal lines show the unit slope.

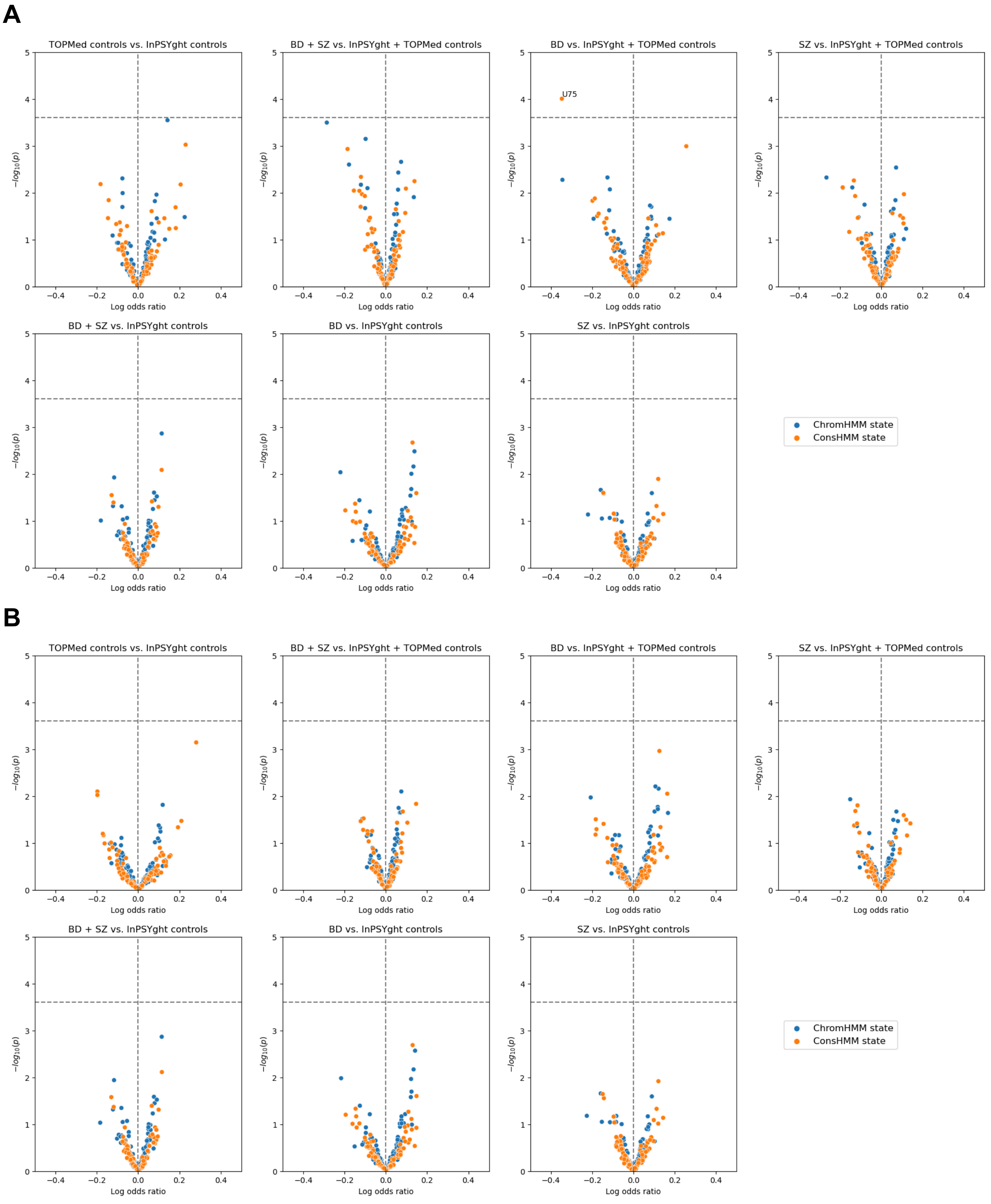


**Figure S12**. Volcano plots of chromatin and conservation state burden test results for different combinations of samples, before (A) and after (B) using sequencing metadata PCs as covariates, with log odds ratio on the x-axis and -log10 p-values on the y-axis. Vertical lines show x=0. Horizontal lines show Bonferroni-based p-value thresholds (p=0.05/200).

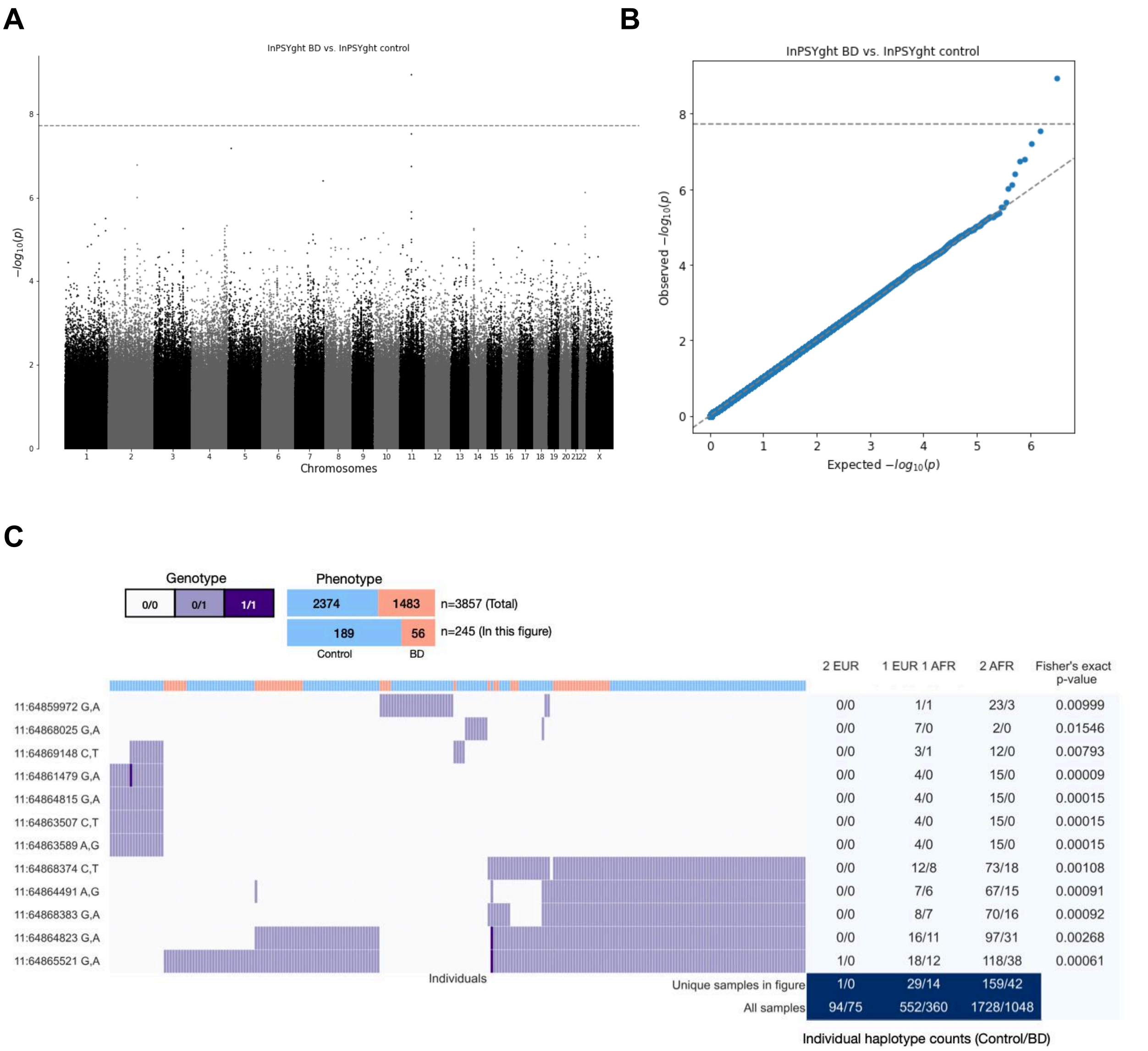


**Figure S13.** Summary of WGScan burden test results for InPSYght BD cases versus controls, with genomic repeat regions removed. BD: bipolar disorder. (**A**) Manhattan plot of all windows being tested; dashed line shows the Bonferroni-corrected p-value threshold based on WGScan's estimated number of effective tests (0.05 / 2,463,399 = $2.0 \times 10^{-8}$). (**B**) Quantile-quantile plot for burden p-values of all windows being tested. (**C**) Sample genotypes for the significant window chr11:64,859,972-64,859,939. The sites with nominally significant Fisher's exact p-values and the samples with at least 1 alternative allele observation among these sites are shown in the heatmap. The table on the right shows the ancestry haplotype distributions for samples having each variant and the Fisher's exact p-values for each variant.

**A**

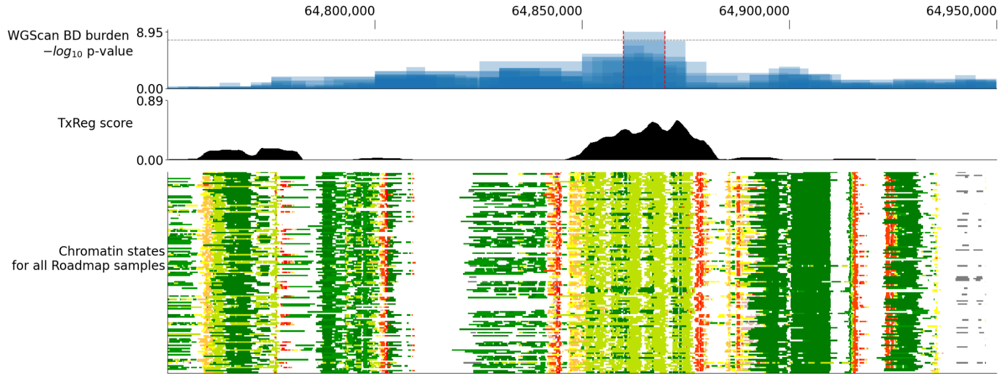


**B**

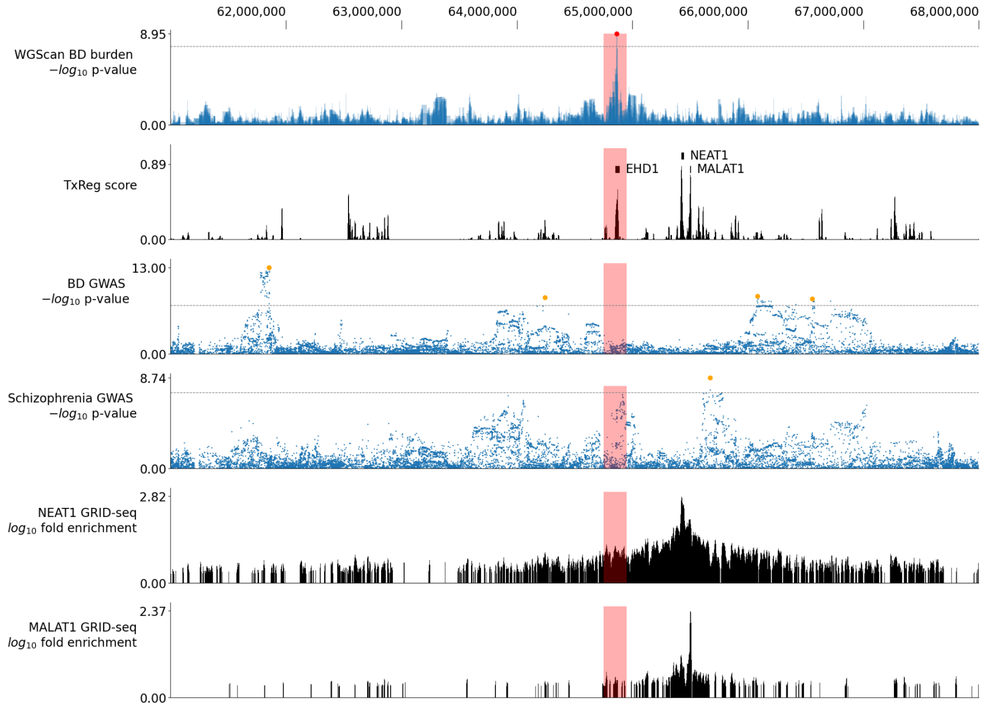


**C**

| State | % of assignments in lead 10kb window | % 10kb windows >= frequency than lead |
|---|---|---|
| TxReg | **44.54** | **0.02** |
| TxEnh5' | 29.61 | 0.08 |
| TxEnhW | 5.45 | 3.38 |
| Tx5' | 16.03 | 4.32 |
| Tx | 2.24 | 4.50 |
| PromD2 | 0.09 | 6.89 |
| Tx3' | 0.13 | 17.40 |
| EnhAF | 0.02 | 23.17 |
| TxWk | 1.15 | 27.46 |
| EnhAc | 0.09 | 34.86 |
| EnhA1 | 0.02 | 35.99 |
| EnhA2 | 0.08 | 37.48 |
| EnhWk2 | 0.19 | 50.73 |

**Figure S14.** Genomic region surrounding the significant window from WGScan BD burden analysis. (**A**) A smaller region surrounding the significant window. Top: windows tested by WGScan shown as bars with height corresponding to their $-\log_{10}$ p-values. The significant window (chr11: 64,859,972-64,869,939) is indicated between the red dashed lines. Grey dashed line indicates the Bonferroni-corrected p-value threshold based on WGScan's estimated number of effective tests ($0.05 / 2,463,399 = 2.0 \times 10^{-8}$). Middle: average TxReg score (computed as the percentage among all 127 Roadmap cell & tissue types that were assigned to the TxReg state) across 200bp bins of a 10kb window around the position. Bottom: chromatin state assignments using the 12-mark 25-state imputation-based ChromHMM annotations[8]. (**B**) A larger region surrounding the significant window (indicated as red boxes). From top to bottom: WGScan windows (same as in (**A**)), TxReg score (same as in (**A**)), GWAS summary statistics from the PGC BD study[5] and the PGC SZ study[6] with genome-wide significant loci shown as orange markers and genome-wide significance threshold shown as grey dashed lines, GRID-seq signal tracks for NEAT1 and MALAT1 on the MM.1S cell line[9]. (**C**) Percentages of 200bp bins assigned to each of the chromatin states (12-mark, 25-state ChromHMM model)[7,8] within the 10kb significant window, aggregated over 127 cell and tissue types, and the percentages of all 10kb windows throughout the genome with greater assignments.

### Supplemental Acknowledgements

#### TOPMed Neurocognitive Working Group

| Last Name | First Name | Affiliation |
|---|---|---|
| Almeida | Marcio | University of Texas Rio Grande Valley School of Medicine |
| Ament | Seth | University of Maryland |
| Ammous | Farah | University of Michigan |
| Arnett | Donna | University of South Carolina |
| Becker | Diane | Johns Hopkins University |
| Bis | Joshua | University of Washington |
| Blue | Elizabeth | University of Washington |
| Boerwinkle | Eric | University of Texas Health at Houston |
| Bressler | Jan | University of Texas Health at Houston |
| Cammann | Davis | University of Nevada Las Vegas |
| Chaar | Dima | University of Michigan |
| Chen | Jingchun | University of Nevada Las Vegas |
| Clarkson-Townsend | Danielle | Brigham & Women's Hospital |
| Coresh | Josef | Johns Hopkins University |
| DeStefano | Anita | Boston University |
| Deters | Kacie | University of California, Los Angeles |

| | | |
|---|---|---|
| Ding | Jingzhong | Wake Forest Baptist Health |
| Fardo | David | University of Kentucky |
| Fitzpatrick | Annette | University of Washington |
| Fornage | Myriam | University of Texas Health at Houston |
| French | Jennifer | University of Maryland |
| Glahn | David | Boston Children's Hospital, Harvard Medical School |
| Gonzalez | Hector | University of California, San Diego |
| Granot-Hershkovitz | Einat | Brigham & Women's Hospital |
| Hanly | Patrick | University of Calgary |
| Hayden | Kathleen | Wake Forest Baptist Health |
| Heckbert | Susan | University of Washington |
| Horvath | Steve | University of California, Los Angeles |
| Hoth | Karin | University of Iowa |
| Hughes | Timothy | Wake Forest Baptist Health |
| Jaiswal | Sidd | Stanford University |
| Jian | Xueqiu | University of Texas Health at San Antonio |
| Katsumata | Yuriko | University of Kentucky |
| Kho | Minjung | University of Michigan |
| Kooperberg | Charles | Fred Hutchinson Cancer Research Center |
| Launer | Lenore | National Institute on Aging |

| | | |
|---|---|---|
| Lin | Honghuang | Boston University |
| Litkowski | Elizabeth | University of Colorado Anschutz Medical Campus |
| Longstreth | Will | University of Washington |
| Mayeux | Richard | Columbia University |
| Mikulla | Julie | National Heart, Lung, and Blood Institute |
| Misra | Biswapriya | Wake Forest Baptist Health |
| Mosley | Thomas | University of Mississippi |
| Nyquist | Paul | Johns Hopkins University |
| O'Connell | Jeff | University of Maryland |
| Olivier | Michael | Wake Forest Baptist Health |
| Panyard | Daniel | Stanford University |
| Peloso | Gina | Boston University |
| Perry | James | University of Maryland |
| Psaty | Bruce | University of Washington |
| Purcell | Shaun | Mass General Brigham |
| Raffield | Laura | University of North Carolina |
| Reiner | Alex | Fred Hutchinson Cancer Research Center, University of Washington |
| Rotter | Jerome | Lundquist Institute |
| Sargurupremraj | Muralidharan | University of Texas Health at San Antonio |

| | | |
|---|---|---|
| Sarnowski | Chloé | Boston University, University of Texas Health at Houston |
| Satizabal | Claudia | Boston University |
| Schellenberg | Gerard | University of Pennsylvania |
| Seo | Jungkyun | University of North Carolina |
| Seshadri | Sudha | University of Texas Health at San Antonio |
| Shade | Lincoln | University of Kentucky |
| Short | Meghan | Boston University |
| Simino | Jeannette | University of Mississippi |
| Smith | Jennifer | University of Michigan |
| Smoller | Sylvia | Albert Einstein College of Medicine |
| Snively | Beverly | Wake Forest Baptist Health |
| Soemedi | Rachel | Brigham & Women's Hospital |
| Sokolow | Sophie | University of California, Los Angeles |
| Thornton | Timothy | University of Washington |
| Vivek | Sithara | University of Minnesota |
| Wood | Alexis | Baylor College of Medicine |
| Yanek | Lisa | Johns Hopkins University |
| Yang | Qiong | Boston University |
| Yu | Miao | University of Michigan |
| Zare | Habil | University of Texas Health at San Antonio |

| | | |
|---|---|---|
| Zhao | Wei | University of Michigan |

#### TOPMed Study-specific Acknowledgements

*Cleveland Family Study – WGS Collaboration (CFS)*

The Cleveland Family Study has been supported in part by National Institutes of Health grants [R01-HL046380, KL2-RR024990, R35-HL135818, and R01-HL113338].

*New Approaches for Empowering Studies of Asthma in Populations of African Descent - Barbados Asthma Genetics Study (BAGS)*

We gratefully acknowledge the contributions of Pissamai and Trevor Maul, Paul Levett, Anselm Hennis, P. Michele Lashley, Raana Naidu, Malcolm Howitt and Timothy Roach, and the numerous health care providers, and community clinics and co-investigators who assisted in the phenotyping and collection of DNA samples, and the families and patients for generously donating DNA samples to the Barbados Asthma Genetics Study (BAGS). Funding for BAGS was provided by National Institutes of Health (NIH) R01HL104608, R01HL087699, and HL104608 S1.

*Genetic Epidemiology of COPD Study (COPDGene)*

The COPDGene study (NCT00608764) is supported by grants from the National Heart, Lung and Blood Institute (NHLBI; U01HL089897 and U01HL089856), by National Institutes of Health contract 75N92023D00011, and by the COPD Foundation through contributions made to an Industry Advisory Committee that has included AstraZeneca,

Bayer Pharmaceuticals, Boehringer-Ingelheim, Genentech, GlaxoSmithKline, Novartis, Pfizer and Sunovion. A full listing of COPDGene investigators can be found at: http://www.copdgene.org/directory

*Multi-Ethnic Study of Atherosclerosis (MESA & MESA AA_CAC)*

Whole genome sequencing (WGS) for the Trans-Omics in Precision Medicine (TOPMed) program was supported by the National Heart, Lung and Blood Institute (NHLBI). WGS for “NHLBI TOPMed: Multi-Ethnic Study of Atherosclerosis (MESA)” (phs001416.v3.p1) was performed at the Broad Institute of MIT and Harvard (3U54HG003067-13S1). The MESA projects are conducted and supported by the National Heart, Lung, and Blood Institute (NHLBI) in collaboration with MESA investigators. Support for the Multi-Ethnic Study of Atherosclerosis (MESA) projects are conducted and supported by the National Heart, Lung, and Blood Institute (NHLBI) in collaboration with MESA investigators. Support for MESA is provided by contracts 75N92020D00001, HHSN268201500003I, N01-HC-95159, 75N92020D00005, N01-HC-95160, 75N92020D00002, N01-HC-95161, 75N92020D00003, N01-HC-95162, 75N92020D00006, N01-HC-95163, 75N92020D00004, N01-HC-95164, 75N92020D00007, N01-HC-95165, N01-HC-95166, N01-HC-95167, N01-HC-95168, N01-HC-95169, UL1-TR-000040, UL1-TR-001079, UL1-TR-001420, UL1TR001881, DK063491, and R01HL105756. The authors thank the other investigators, the staff, and the participants of the MESA study for their valuable contributions. A full list of participating MESA investigators and institutes can be found at http://www.mesa-nhlbi.org.

*Atherosclerosis Risk in Communities study (ARIC VTE)*

The Atherosclerosis Risk in Communities study has been funded in whole or in part with Federal funds from the National Heart, Lung, and Blood Institute, National Institutes of Health, Department of Health and Human Services, under Contract nos. (75N92022D00001, 75N92022D00002, 75N92022D00003, 75N92022D00004, 75N92022D00005). The authors thank the staff and participants of the ARIC study for their important contributions.

Whole genome sequencing (WGS) for the Trans-Omics in Precision Medicine (TOPMed) program was supported by the National Heart, Lung and Blood Institute (NHLBI). WGS for “NHLBI TOPMed: Atherosclerosis Risk in Communities (ARIC)” (phs001211) was performed at the Baylor College of Medicine Human Genome Sequencing Center (HHSN268201500015C and 3U54HG003273-12S2) and the Broad Institute for MIT and Harvard (3R01HL092577- 06S1).

*Women’s Health Initiative (WHI)*

The WHI program is funded by the National Heart, Lung, and Blood Institute, National Institutes of Health, U.S. Department of Health and Human Services through contracts 75N92021D00001, 75N92021D00002, 75N92021D00003, 75N92021D00004, 75N92021D00005.

*Hypertension Genetic Epidemiology Network (HyperGen)*

The HyperGEN Study is part of the National Heart, Lung, and Blood Institute (NHLBI) Family Blood Pressure Program; collection of the data represented here was supported by grants U01 HL054472 (MN Lab), U01 HL054473 (DCC), U01 HL054495 (AL FC), and U01 HL054509 (NC FC). The HyperGEN: Genetics of Left Ventricular Hypertrophy Study was supported by NHLBI grant R01 HL055673 with whole-genome sequencing made possible by supplement -18S1.

*Genetics of Sarcoidosis in African Americans (Sarcoidosis)*

National Institutes of Health (R01HL113326, P30 GM110766-01)

*Mount Sinai BioMe Biobank (BioMe)*

The Mount Sinai BioMe Biobank has been supported by The Andrea and Charles Bronfman Philanthropies and in part by Federal funds from the NHLBI and NHGRI (U01HG00638001; U01HG007417; X01HL134588). We thank all participants in the Mount Sinai Biobank. We also thank all our recruiters who have assisted and continue to assist in data collection and management and are grateful for the computational resources and staff expertise provided by Scientific Computing at the Icahn School of Medicine at Mount Sinai.

*Coronary Artery Risk Development in Young Adults (CARDIA)*

The Coronary Artery Risk Development in Young Adults Study (CARDIA) is conducted and supported by the National Heart, Lung, and Blood Institute (NHLBI) in collaboration with the University of Alabama at Birmingham (75N92023D00002 & 75N92023D00005), Northwestern University (75N92023D00004), University of Minnesota (75N92023D00006), and Kaiser Foundation Research Institute (75N92023D00003). CARDIA was also partially supported by the Intramural Research Program of the National Institute on Aging (NIA) and an intra-agency agreement between NIA and NHLBI (AG0005).

#### Supplemental References

1. Wei, S., Xu, Y., Shi, H., Wong, S.-H., Han, W., Talbot, K., Hong, W., and Ong, W.-Y. (2010). EHD1 is a synaptic protein that modulates exocytosis through binding to snapin. Mol. Cell. Neurosci. *45*, 418–429. https://doi.org/10.1016/j.mcn.2010.07.014.
2. Yap, C.C.C., Lasiecka, Z.M., Caplan, S., and Winckler, B. (2010). Alterations of EHD1/EHD4 Protein Levels Interfere with L1/NgCAM Endocytosis in Neurons and Disrupt Axonal Targeting. J. Neurosci. *30*, 6646–6657. https://doi.org/10.1523/JNEUROSCI.5428-09.2010.
3. Kataoka, M., Matoba, N., Sawada, T., Kazuno, A.-A., Ishiwata, M., Fujii, K., Matsuo, K., Takata, A., and Kato, T. (2016). Exome sequencing for bipolar disorder points to roles of de novo loss-of-function and protein-altering mutations. Mol. Psychiatry *21*, 885–893. https://doi.org/10.1038/mp.2016.69.
4. Nakamura, T., Nakajima, K., Kobayashi, Y., Itohara, S., Kasahara, T., Tsuboi, T., and Kato, T. (2021). Functional and behavioral effects of de novo mutations in calcium-related genes in patients with bipolar disorder. Hum. Mol. Genet. *30*, 1851–1862. https://doi.org/10.1093/hmg/ddab152.
5. Mullins, N., Forstner, A.J., O’Connell, K.S., Coombes, B., Coleman, J.R.I., Qiao, Z., Als, T.D., Bigdeli, T.B., Børte, S., Bryois, J., et al. (2021). Genome-wide association study of more than 40,000 bipolar disorder cases provides new insights into the underlying biology. Nat. Genet. *53*, 817–829. https://doi.org/10.1038/s41588-021-00857-4.
6. Trubetskoy, V., Pardiñas, A.F., Qi, T., Panagiotaropoulou, G., Awasthi, S., Bigdeli, T.B., Bryois, J., Chen, C.-Y., Dennison, C.A., Hall, L.S., et al. (2022). Mapping genomic loci implicates genes and synaptic biology in schizophrenia. Nature *604*, 502–508. https://doi.org/10.1038/s41586-022-04434-5.
7. Ernst, J., and Kellis, M. (2012). ChromHMM: automating chromatin-state discovery and characterization. Nat. Methods *9*, 215–216. https://doi.org/10.1038/nmeth.1906.
8. Ernst, J., and Kellis, M. (2015). Large-scale imputation of epigenomic datasets for systematic annotation of diverse human tissues. Nat. Biotechnol. *33*, 364–376. https://doi.org/10.1038/nbt.3157.
9. Li, X., Zhou, B., Chen, L., Gou, L.-T., Li, H., and Fu, X.-D. (2017). GRID-seq reveals the global RNA–chromatin interactome. Nat. Biotechnol. *35*, 940–950. https://doi.org/10.1038/nbt.3968.
10. Yang, S., Lim, K.-H., Kim, S.-H., and Joo, J.-Y. (2021). Molecular landscape of long

noncoding RNAs in brain disorders. Mol. Psychiatry *26*, 1060–1074. https://doi.org/10.1038/s41380-020-00947-5.
11. Srinivas, T., Mathias, C., Oliveira-Mateos, C., and Guil, S. (2023). Roles of lncRNAs in brain development and pathogenesis: Emerging therapeutic opportunities. Mol. Ther. *31*, 1550–1561. https://doi.org/10.1016/j.ymthe.2023.02.008.
12. Katsel, P., Roussos, P., Fam, P., Khan, S., Tan, W., Hirose, T., Nakagawa, S., Pletnikov, M.V., and Haroutunian, V. (2019). The expression of long noncoding RNA NEAT1 is reduced in schizophrenia and modulates oligodendrocytes transcription. Npj Schizophr. *5*, 3. https://doi.org/10.1038/s41537-019-0071-2.
13. Shirvani Farsani, Z., Zahirodin, A., Ghaderian, S.M.H., Shams, J., and Naghavi Gargari, B. (2020). The role of long non-coding RNA MALAT1 in patients with bipolar disorder. Metab. Brain Dis. *35*, 1077–1083. https://doi.org/10.1007/s11011-020-00580-9.
